# Mechanisms and attitudes in responsive health care for forced migrant communities. A qualitative study of transnational practice

**DOI:** 10.1101/2024.11.29.24309172

**Authors:** Amy Robinson, Ziaur Rahman A. Khan, Kofi Broadhurst, Laura Nellums, Gisela Renolds, Bayan Payman, Andrew Smith

## Abstract

**Objectives:** To understand the opportunities and practices that can support responsive health care for forced migrant communities.

**Design:** Qualitative study of five transnational case examples of services actively working to improve access and experiences of care for forced migrant communities. One strand of the MORRA Study.

**Setting:** Five services (Australia, Belgium, UK) providing a range of care (primary care, health advocacy, education and support, holistic health screening, care planning/coordination, transcultural mental health care). Delivered through state and not-for-profit structures in initial and contingency accommodation sites, health clinics, and community spaces. Data collection took place between July and October 2022.

**Participants:** 47 participants including forced migrants using or having used one of the five services, service leads, clinical and non-clinical workers (paid and volunteer), interpreters, and service partners. Services supported recruitment of a crude representative sample of worker roles and service users/clients. Participants were required to speak one of nine languages for which we had translated study materials.

**Main outcome measures:** Experiences, practices, knowledges, skills and attributes of workers; experiences of forced migrants engaging in services.

**Results:** Services showed a willingness to innovate and work outside existing practice and organisational structures, including a ‘micro-flexibility’ in their interactions with patients, and through the creation of safe spaces that encouraged trust in providers. Other positive behaviours included: engaging in intercultural exchange; facilitating the connection of people with their cultural sphere (e.g. nationality, language); and a reflexive attitude to the individual and their broader circumstances. Social and political structures can diminish these efforts.

**Conclusions:** Environments that enable good health and support forced migrants to live lives of meaning are vital components of responsive care. This requires flexibility and reflexivity in practice, intercultural exchange, humility, and a commitment to communication. A broader range of caring practitioners can, and should, through intentional and interconnected communities of care, contribute to the health care of forced migrants. Opening up health care systems to include other state actors such as teachers and settlement workers and a range of non-state actors that should include community leaders and peers and private players is a key step in this process. Future work should focus on the health and health service implications of immigration practices; the inclusion of peers in a range of health care roles; alliance-building across unlikely collaborators and the embedding of intercultural exchange in practice. Findings of this study are supported by our systematic review (publication forthcoming).

**Strengths and limitations of this study:**

- We engaged with critical perceptions of care from across a range of international jurisdictions, community and health service contexts, and clinical and non-clinical professionalisms.
- A diverse and contrasting research team, including a multilingual community researcher, NHS clinicians, voluntary sector advocacy services, and academics, brought contrasting perspectives and backgrounds and broadened reflections.
- Unexpected restrictions (predominantly service pressures and restrictions placed by service hosts – accommodation providers) meant we engaged only a small number of service users and only with service users from some services.

## Introduction

The human repercussions of forced displacement are complex and diverse. Adversities that begin before displacement and continue throughout flight are often replicated and upheld in arrival and settlement contexts where factors associated with language, culture, knowledge, gender, race, and citizenship^1,2^ can intersect and continue to influence and shape health, health care and settlement experiences.^2–5^

Responding to these complexities and recognising their influence on a person’s physical, mental and psychosocial health is a widely agreed priority.^2–10^ Yet, though legal structures and heavy evidence provide an ethical framework on which to recognise rights to care and the social and material conditions for good health,^11^ including accessible and acceptable public systems, there are profound disconnects between receiving public systems and the needs of forcibly displaced populations. ^9,12–15^

From skill gaps amongst care providers when operating in cross-cultural interactions,^12,14,16^ poor communication^9^ and failure to recognise fundamental rights to care,^17–20^ to low levels of understanding around the health risks and experiences of displaced communtities,^7^ a range of factors can undermine the accessibility and quality of health care provision.^17–19,21^ Systemic and structural factors including what are generally recognised as inflexibilities in public systems of care further limit practitioners’ capacities to respond to the often additional and comorbid health needs that forced migrants present in comparison to local populations.^4,17,22,23^ Equally, through their explicit delivery or interpretation,^24^ public policies of *hostility* can have direct and more nuanced impacts on health and access to care^25–27^ which, through establishing hierarchies of deservingness,^28^ can perform as everyday bordering in public sector health care settings ^27,29–32^ and play a determinant role in health and help-seeking.^10,33^

There is also limited attention given to securing knowledge for new arrivals around both rights to care^34^ and navigation of local health systems.^10,35^ Combined with altered social roles, socio-cultural and ethno-religious isolation and the challenges of adapting to a new dominant culture^36^ repercussions can include a reduced sense of self, accumulative stress,^37,38^ and a disconnection from trusted (and often traditional sources of) support, further contributing to a cycle of poor health that is both fed by and feeds low social participation.^39,40^

Clarity around how to adopt refugee- and migrant-sensitive health systems^17,41^ and demonstrate solutions to some of the health care barriers and poor experiences in displacement contexts^42^ remains a gap. It is this absence that MORRA (NIHR132961/ CRD42021271464), a mixed-methods study combining participatory aspects, community engagement, a mixed-methods systematic review and the qualitative analysis of five contemporary case studies, sought to address. Reported here are the qualitative case studies. The systematic review and an overall synthesis of findings will be published in a forthcoming report.

### Theoretical Framework

While acknowledging that forced migrants are a heterogeneous group, we use this as a collective term to refer to people who have been forcibly displaced and cannot return home safely. We include here people who may be described as asylum seekers; refused asylum seekers; refugees; undocumented people; and unaccompanied -asylum seekers, -children, or -young people. We do this while also acknowledging the risk of arbitrary categorisation, the absence of agreement on where the line between voluntary and forced migration may be drawn,^43^ and that each of these classifications are recognised by nation states and others in different ways with ideas of deservingness and associated rights, with consequential influence on access to care and broader adversities faced.

Different frames of reference encourage us to think about broader conditions and contexts when considering health care for forced migrants. Notably, the responsibility of receiving societies to shape refugee-integration opportunities,^44^ and ecological^45^ and social justice^46^ perspectives that place attention on the contextual realities in which a person is located. As we understand it, these each place life situations and broader environments as central influencing factors in how we might consider the assessment and identification of someone’s needs, and importantly where health care responses are best located for people with complex migratory and post migratory situations.^47,48^ Incorporating intersectionality into research design, can, as Windsong ^49^ suggests, help us to democratise public health and encourage discussion that brings to the surface oppressions that potentially damage health care interactions for those at the intersection of different marginalised experiences, including race, gender and (dis)ability^50^ encouraging attentiveness to specific experiences of advantage and disadvantage.^51^

It is through these overlapping frameworks that we approach this research: specifically locating our interest in a frame that acknowledges the systematic marginalisation, particularly of people without formal refugee status^32^ and the value of a health response that considers the multiplicity and multidirectional influences affecting health, wellbeing and the capacity to engage with and receive good care.

This considered, we sought to learn from different jurisdictions and models of care that were actively taking steps to consider these factors. We used a case study methodology to understand possibilities and practices and located these within the different local knowledges, experiences, skills, and attributes of workers alongside the experiences of those engaging in services while also taking account of the broader institutional, political, and community contexts in which this all takes place. Semi-structured interviews with providers, collaborators and service users allowed us to gain insight about everyday experiences and practices, the subjective value, essential aspects and contributions individuals attributed to each service, and the further developments in care that were seen to be important. Visits and observations broadened our understanding of the local contexts, providing the opportunity to acquire the tacit knowledge that can be drawn from observing workers personal qualities, the range of interactions taking place within a given space^52^ and the informal reflexive perceptions and insights between participants and researcher,^53^ bringing a vital aspect to our enquiry.^54,55^

### Objectives

Characterising responsive care as that which actively seeks to notice and appropriately respond to the particular needs (and context) of a given individual we sought to identify current examples of care that were taking active steps to improve access to and experiences of health care for forced migrants. We aimed to identify promising and divergent practices and to understand the explicit mechanisms being used to improve health care access, to identify providers’ knowledges, relationships, experiences, skills and attributes, and understand these as part of the broader local context and experiences of those engaging with the service.

Using Saurman’s^56^ interpretation of access (and consideration to the interconnected domains of: physical access, availability, acceptability, affordability, design and usability, and awareness) in conjunction with attention to health agency as to whether people have the tools and right conditions to utilise those domains

## Methods

### Study design

This qualitative study drew on ethnographic methods including semi-structured interviews, observation and informal conversation.

To support recruitment, we used academic, clinical, refugee, and migrant networks, contacted a range of national and international migration and refugee agencies, conducted web-based searches, used social media, and identified possible services when screening studies in our associated review. We sought services operating in any country and used criteria based on variability in health care opportunity, health need of focus, delivery location, patient group, organisational context, and theoretical or ideological approach to agree case study selection.

We relied on a service contact to assist with the recruitment of previous or current service users (encouraging participants with different identity characteristics and backgrounds such as immigration status, local language skills, time in country, health needs, gender, lone individuals/families) and workers and organisation partners (aiming for representation of different roles).

Authors AR and ZK conducted group and individual interviews, site visits, service observations and informal conversations with workers between July and October 2022. Using the criteria above, we used criterion-based purposive sampling,^57^ aiming to recruit six individuals (including service users, workers, partners) from across each service. Given service location and capacity, a wide variation in the staff make-up of each service, and research team capacity, we made a pragmatic decision to accept some variation in recruitment. Engagement with services loosely equated to two-days per case study for site visits, observation, informal conversation and face-to-face interviews and one additional day per case study for remote video interviews. Most interviews lasted one hour. Interviews with service users were conducted in Dari or Pashto by ZK, or with interpreters.

Three staff interviews involved interpreters. All participants provided informed consent and interview and observation guides (see supplementary files) were used during conversations and observations of practice. Most interviews were recorded except where two service users requested otherwise (we did not push for an explanation but both individuals were quick and firm in their objection). Written notes were made during these conversations, which were discussed for accuracy between AR and ZK following the interview and used in analysis in the same way as the recorded and transcribed interviews. Field notes were made following site visits and observations. Reflective conversations took place between AR and ZK immediately following site visits and again after all field notes for each visit were notated. We collected any available service information sheets and published evaluations from services.

### Research practice for cross-cultural settings

Study materials and interview and observation guides were developed iteratively with the multidisciplinary project team (including people with lived experience) with final versions (pre-translation) proofread by two people with refugee backgrounds. Forced migrant participant materials gave consideration to avoiding distress and re-traumatisation and we did not encourage discussion of displacement experiences or individual health conditions. We emphasised information shared would remain anonymous and would not be shared beyond the research team. Service user materials were translated into nine languages commonly spoken by users of the five services (Amharic, Arabic, Dari, Farsi, Kurdish Sorani, Oromo, Pashto, Spanish, and Tigrinya). This was undertaken by professional translators who were native speakers and checked by a second native speaker. Despite this, later feedback from native speakers flagged some of these materials as wooden and lacking coherence, reiterating the challenges in multilingual communication. We were flexible in the manner in which we talked with service users: individually, in groups, face to face or via WhatsApp video. Though we were using the collaboratively developed interview and observation guides, we allowed conversations to diverge in directions important to participants, remaining conscious of the power imbalances between researchers and participants. Although we identify the case study services, given the small number of participants involved we do not tend to attribute participant narrative to a service or role to protect confidentiality and anonymity.

### Analysis of results

Authors (AR, KB) anonymised and transcribed a verbatim account of each interview using colloquial language, broken language and grammar as spoken. Transcripts were read by AR and an initial coding frame was developed through an inductive and ‘bottom-up’ data-driven approach. This was discussed and amended with ZK before being finalised. Coding was undertaken by AR using Nvivo12 software and included the coding of fieldnotes and service documents. Guided by our research aims and following principles of thematic analysis^58^ coded extracts were then developed iteratively into broad themes. A reflective discussion was undertaken with ZK to agree final themes and findings. This analysis took an essentialist, realist approach,^59^ reporting experiences, focusing on the meanings in the data and as Braun and Clarke^58^ indicate in their description of the method, reporting the reality of participants that could be interpreted from the conversations conducted and observations of practice and contexts.^59^ We gave the five participating services the opportunity to check initial findings for accuracy and resonance.^60–62^

### Researcher positionality

ZK is a multilingual (Pashto, Dari, English with some Arabic and Farsi) reintegration caseworker working with asylum-seeker and refugee communities. He has an educational background in Agriculture, Economics and Planning, and Business Administration (MBA). ZKs shared language with some participants was well received and brought a degree of “lived familiarity”^63^ facilitating a trusting and relaxed interaction. ZK’s role as a health advocate and integration officer working with people with refugee backgrounds added a further direction of interest in conversations with service providers. AR is a mixed methods researcher with more than ten years’ experience undertaking qualitative health research within communities and health care settings in the UK. Her training lies in conflict resolution (regional/state) and policy evaluation. She has trained with the Children and War Foundation in recognising and responding to the psychosocial impacts of experiences of war and is interested in rights to recognition in health care interactions. AR approached the work as an ‘insider’ of the UK health service but has wide experience conducting qualitative research within and across different sectors often relating to access to care and is involved, as a volunteer, in local refugee responses. Depending on the context and participants interviewed we moved in and out of different roles. Though we acknowledge that all these positions bring with them subjectivities we saw them as strengths bringing a degree of balance to the research. KB is a PhD candidate in the area of critical sociology focusing on imperial memory, decolonial thought and white identity in Britain.

Ethical approval was granted by the Health Research Authority London Bridge Research Ethics Committee, 1^st^ June 2022, ref: 22/PR0603.

### Patient and public involvement

We actively implemented an inclusive project approach that attempted to soften the boundaries of research as an externalist practice.^63^ Our project team was multisectoral and included people with expiriences of forced migration, refugee and asylum-seeker support services, health and other public service providers, and academics. This brought a diversity of perspectives to decisions and practice; helped to emphasise the importance of clarity in communication which needed to be succinct and framed for different audiences; and helped to clarify expectations we might have of different participants and the lines of enquiry we should follow. We sought additional input from others with lived experience in relation to planning and co-facilitation of community workshops and planning content and reviewing of participant materials.

We delivered a range of community workshops, conversations and events with forced migrants (46 individuals with experiences of forced displacement were involved across four sessions) and the voluntary, public and private sector. These were intended to support the validity and relevance of our research in both the needs of forced migrant communities and those of caring practitioners in the field, and was an attempt to reach some of those who may be able to utilise our learning. A summary of these events is reported in a forthcoming report.

Forced migrant community members who participated in the research or attended workshops were provided vouchers to acknowledge their contribution.

## Results

Five services were chosen from a longlist of 130 providers. We visited three services (UK only) and observed service activities over five-days. Observations included reception spaces, social spaces, drop-in activities, and clinical appointments. We undertook 24 one-to-one interviews in person (12) and over Microsoft Teams (12) and five in-person group-interviews which included a total of 11 individuals. We had informal conversations with 12 workers.

Participants included: clients and services users (most were male and lone adults covering a range of age groups and from the MENA region, sub-Saharan Africa, and South East Asia; all had literacy in their native language), service managers/directors, health advocates, interpreters, office managers, wellbeing project leads and staff, social prescribers, reception staff, nurses (migrant health, refugee health and inclusion health nurses, chief nurses), mental health workers (leads, ethnotherapists, psychologists, psychiatrists), clinical consultants, GPs, health care assistants, network managers, communications and fundraising staff, project volunteers, and project partners. We do not report specific numbers due to the small number of individuals from each group.

### Five case examples

Key details relating to our five case examples can be found in supplementary material. All services worked with children, adults and families. Four services worked predominantly with people who were within two years of arriving within the local country, while one service worked with people at any point in their settlement journey. Services stressed a needs-led approach with people often never formally discharged from care and told clearly how to re-engage where needed.

Three services were based in the UK, one was based across Belgium, and one in Victoria, Australia. Services were generally funded through a combination of local and national state health commissioning, regional/global migration funds, charitable trusts and foundations, as well as some informal sources (faith organisations and private donors). All services were free at point of access.

Delivery settings included initial and contingency accommodation (4 services), health clinics (4 services), and community centres or spaces, including schools (3 services). Two services were co-located with community, wellbeing and social support services and three services described occasional or initial home visits. A summary of offered provision is detailed in supplementary material. Services drew on a range of clinical, non-clinical health workers, and other roles such as social workers and intercultural mediators. Some services included volunteers and peers. Full details are provided in supplementary material. Services put substantial effort into supporting all staff.

All services had a central commitment to good communication, including mechanisms for keeping a record of the languages spoken and whether people could read and write in their own language. These details were included in external referrals. All clinical services had access to telephone language services. Two services worked almost entirely with professional, in-person interpreters, one entirely with informal in-person interpreters, one service used telephone interpreters only, and a final service engaged a combination of all three. Some services had in-house or pseudo in-house block-booked in-person interpreters. Interpreters were often valued as part of the team and well-paid to acknowledge their skill and contribution.

Printed materials were sometimes produced in clients’ common languages and some services utilised simple colour printed illustrations with no text for patient messaging.

### Communities of care: Themes and findings

Our interviews, conversations and observations highlighted a set of largely shared attitudes, priorities and capacities seen as important to working with forced migrant communities. These are summarised in Figure 1 and discussed below. They should be seen as interconnecting and overarching, and not the totality of practice.

**Figure 1.** Communities of care

### Attitudes of care

Attitudes of care were held together by three interconnected themes.

#### Innovating and adapting

Models of care required constant re-invention and the navigation of unpredictability as services responded to changing policy environments, local contexts and the changing needs of patients and clients.

“A case of testing something… if it doesn’t work you trial, you trial, you test it… it doesn’t work, you go back to the drawing board, and you look at different ways.”

Appointment systems were one example, where services tested out: flexibility for walk-ins “because some people do just need seeing there and then”; primary care triage that actively allowed for a social or a clinical response; and flexing on length of appointments.

There was an increasing inclusion of social workers and care coordinators in one nurse-led team to “get a handle” on the social support needs of increasing numbers of people seeking asylum. There was common reference to a “duty of care” to see people as soon as they were made aware of new patients, requiring constant navigation of the ebbs and flows of patient numbers, noticing and responding to emerging needs, and evolving practice and ways of working: “This is our problem… how can we think about it, how can we be more effective?”

Where services were providing health care to residents in contingency accommodation there was a continuous process of identifying new locations, rethinking practices, negotiating access and local collaborations, and accepting a heavy resource commitment to operations and relationships.

> *“What became clear, very quickly, was that you couldn’t set it up in one way and stick to it because the need kept moving and as soon as a hotel shut something else opened and every patch had a different infrastructure… what we learned was that we needed to be really quick… we needed to set up where we were needed and in the way that we were needed.”*

Navigating working relationships with accommodation providers was feasible but also combined with a “general hostility”: accommodation staff were sometimes “obstructive”, care providers were “up against it to get in to help people” and at times were “turned away by security”. Communication challenges, exacerbated by high staff turnover within accommodation contractors, was seen as a key factor.

Care within these settings also demanded flexibility as to where workers were prepared to do their work, often adapting from one day to the next. Care often delivered “in a little corner” of a communal “lounge” (more corridors than clinics), in rooms (like cupboards) with no windows, no air, and no space, or in a staff room, with security sleeping as bloods were taken. Though there was always effort to make someone feel comfortable, there was still concern at the inability to “control the environment” and that spaces were not always appropriate or conducive to the disclosure of health concerns.

> *“Language Line on loud speaker… being very close to the dining room and then all the stuff going on there getting prepped for dinner and then somebody coming by with a hoover and it was just very, very noisy and the person I was with just got very overwhelmed with it all and very sort of frustrated, he couldn’t sort of tell me, couldn’t hear the interpreter, I couldn’t hear what the interpreter was saying.”*

Worker’s roles had a mobility to them. Care was often being delivered outside of the typical boundaries of usual practice (“there are lots of things that we will be more flexible about, than we might in other services”). It saw psychologists and nurses describe, “sometimes we feel like we are lawyers, sometimes we’re social workers…”, and there was talk of being in a position to act as entrepreneurs, to see challenges and create opportunities to develop a response. There was recognition that these attitudes were made possible first by institutional structures. Though one service described a large health agency as “the opposite of blinkered”, services generally talked about the “possibility” and “focus” that came with sitting outside the “restrictive agendas” of certain agencies.

> *“We’re not constrained by red tape, so something like this, which is a new project, let’s recruit staff for it, let’s have a look at skill mix. Let’s get the money to support* them*. Let’s try it. If it doesn’t work, we’ll try it a different way, that’s the kind of thinking. And there is that flexibility to say, look, actually, there is an unmet need here, what can we do here… it is really, really refreshing.”*

The active involvement of senior leadership in delivery of most services was seen as enabling this. As one provider put it,

> *“[They] are also* invested*… things happen and things happen a lot more easily and a lot more quickly and innovation does turn into… you know… a real tangible service for people. We’re just like… that’s a good idea let’s make it happen…”*

The specialist nature of each service also appeared to help: “Everybody’s head is in the same place… and it’s easier to get my head around the ideas that people have because it’s my world as well.” Workers were “free… to be looking for what the problems are”, “to notice”, to have ideas and “still discovering together” the best ways of delivering care. This was performed through “a different kind of culture”, a mentality of “creativity”, more a “vocation” than a job: “I don’t think it would suit someone who wanted to go in and work by the book.”

#### Flexing for patient need

There was flexibility too in the micro-interactions with patients and clients, which saw women’s groups shaped by women’s interests; workers in a position to enact fluid practices of referral (“If someone comes to one of our groups and just expresses a support need then we’ll pick that up”); and health drop-ins where anything goes (e.g. helping someone to understand a referral letter for the hospital, linking a volunteer with someone trying to find the GP practice, sourcing bikes, providing shoes (*most residents with only flipflops on their feet*)).

Patient engagement was a priority. Word of mouth had proved best for one service engaging hotel residents: “They need to hear that it’s happening…” and here volunteers were recruited “to get them spreading out the word”. An arrangement in another service allowed for young children to be cared for by the health care assistant, with a door propped open for everyone’s peace of mind, but with enough privacy for a parent to discuss more sensitive issues.

There was heavy reflection on why someone might not attend an appointment, recognising that trauma may not make it “easy to come” or *will* make it easy “to forget”. It was described that someone might arrive at their clinic (rather than the hospital) because it makes sense to go to the one that you know and not the one described in the “letter that you don’t understand”. It was often described that people simply cannot get there: “I didn’t have the money”, a “taxi doesn’t come, or it comes late and they miss the appointment”.

To support patient engagement, there was reference to volunteers in hotels knocking on clients’ doors “encouraging” people to attend a planned appointment, text message reminders a day before an appointment, and phone calls prompting or checking that someone was planning to attend or was on their way, clarifying that “it’s okay to be a bit late”, “we’ll fit you in”.

> *“It depends on the patient really. So, some patients, if it gets to 50 minutes late and it’s something really minor, we’ll be like ‘no sorry you need to play by the rules’ if its half an hour… some people will let way for half an hour, some people if they turn up at all we’re just grateful. It depends on how vulnerable they are and so there’s always a bit of flexibility.”*

There was an emphasis on “finding out why” an appointment was missed: “The first thing we do is, we pick up the phone”. Services talked about re-offering appointments, putting in place support, such as help with navigation, telling the time, and peer advocates emphasising the benefits of the offered care: “We will… talk to them and make them understand how useful that is, help them see what they’re missing”. There was occasional reimbursement of travel costs for some appointments but a view that, with a “bit of perseverance” or support, people will subsequently attend.

One psychologist explained, “well for us it’s like, okay, they’re not here, we’re going to go out, we’re going to mobilise”, explaining that stepping outside the clinic was a culture sometimes difficult for new colleagues. Similarly, other services described making home visits for first contact, or recognised that on occasion, for a mother alone with four children, the clinic should go to them.

#### Trust

Trust was a theme threading through many conversations. There was recognition of “huge amounts of fear” with people “worried about disclosing information” and “getting a bit alarmed” by medical consent forms. This was often attributed to not knowing “who to trust”, to experiences of racism and discrimination, and to fear of government institutions. There was sometimes reluctance around and refusal of catch-up vaccinations.

> *“People are worried about being tagged with the vaccines… a colleague, from a different community… pointed out that… they feel like the Home Office might track them, or something… I suppose it’s kind of having that conversation.”*

Workers acknowledged that there was no “expectation [for people] to be open and cooperative after one meeting”, with one explaining; “in the initial contact they wouldn’t trust me, and I don’t blame them… that’s why my first contact, I don’t ask any sort of gruelling questions, it’s just about getting to know them”. Services focused on explaining information and processes and working really slowly, providing “active reassurance that there are no ties” with the “Home Office”, with “the police”… “being really clear about who we are…” and that nothing is “compulsory”.

Creating trusted spaces included the service “welcome”. Reception spaces were intentionally designed with language provision, peer support, and a conscious drive to get it right: “the welcome to the surgery, how to register, how to book an appointment, that is actually one of the key things”. This included seeing receptionists “as part of the team” who might themselves refer people and tell patients about the social and wellbeing support available. It might just be “having a welcoming smile”… “saying ‘welcome Mr and Mrs, have a seat, do you want some coffee? Want some tea? Welcome to […], smile…”. It might be a first interaction in which a worker acknowledges someone’s situation, shows some familiarity with someone’s home country, a shared connection over a local dish, *providing a moment of happy reflection*, workers making efforts to learn “a handful of phrases to support relationship-building”.

> *“Maybe, what also helps is that… when I speak Arabic to people it opens up as well. So, it helps, and I’ve never been to Afghanistan, but I also ask the interpreter, ‘*teach *me some words’ so I can at least greet a person in their mother tongue and that’s… people are smiling andare saying you know ‘do you know more?’ and I’m like ‘no, no, no, […] is difficult’, and then we talk about languages, how hard it is.”*

Mental health was seen as an important, yet taboo topic; “talking about your problems outside of your circle, it’s just not done… you would be completely rejected by your community”. Here there was emphasis on the importance of a “good” relationship; with that “we have already gone half of the way”, slowing things down, and as one worker put it, “there is no limitation for being patient with patients”.

Building on one-to-one emotional support and doing small practical things to build trust was common: “By showing people their rights, for example, you prove that you are worth the while to listen to and then you can start opening up and relating”, one mental health worker, explained. Several staff members talked about concentrating on peoples’ hobbies and interests, “slowly, slowly and then they will come to you and they’ll tell you ‘actually I’m struggling with this and that… can you support me?’” Face-to-face contact was seen as important, “to build that rapport” and avoid patients’ fears of engaging online: as one interpreter explained, “He feels everybody is hearing everything… he does not feel safe to say everything”. The need to avoid barriers such as “a demand for someone’s ID, their address” while completing GP registration, “not taking photocopies” was recognized among staff. A flexibility with appointment times, frequency and location, and valuing the investment of someone’s time as important and part of the “relationship of trust” were further mechanisms, decided mutually. Outreach practices were seen here as a facilitator, closing down some of the barriers and equalising relations between individuals, families, state structures and clinical services:

> *“So when we were assisting the teacher in the conversation with the parents on how to express the problems that a child had in school this was much more like trust* building*, and not, okay, being sent by the teacher to the hospital as it happens normally because the teacher says, ‘I suppose you have a psychiatric problem’, these kind of referrals do not work. So we do the other way, you need to go there”*.

Building trust and rapport was explained by one worker as “the most important thing in our job… to make that connection to provide the situation that they can trust us”, it was a process, it took time, it could take months, and “some people remained suspicious”.

### Navigating variabilities in perceptions and needs

#### Intercultural exchange

Navigating different viewpoints and being aware of the complexity of an intercultural exchange was seen as important in creating and sustaining patient engagement. This required “recognition of different health beliefs” and different interpretations, for example, of “behaviour”. There was a need to resist “ethnocentrism” and institutional cultures, several workers acknowledging the “white majority” for whom health services were designed and the “conditioning” of their practice training which they felt they had to resist. This saw a preparedness on the part of workers (and managers) to reflect upon their own values and motives,

> *“I think it’s… stop pretending that everyone that works in the health service understands the needs of these populations… they don’t. Be humble, listen to the patients, start including peers in the decision-making process from commissioning down”*.

There was recognition of boundaries between normality and pathology and symptoms and syndromes that might be understood and explained differently among different cultures. There was some acknowledging of somatic expressions of mental health and of seeking to understand someone’s own interpretation of what was going on, “okay… what you went through… what did it do for the body?”

There was reference to adapting classical therapeutic techniques, one provider talking about using only the physical aspects of EMDR therapy because cognition and emotion were seen to be less present in their patients: “Within our refugee conception of themselves, it’s much more the body…. war trauma… an unaccompanied minor… violated… it’s too heavy for words but his body will express it”. Here, the same provider explained that often more physical practices such as yoga and stress relief exercises might be the best approach.

Explicit intercultural dialogue was described as part of the outreach methodology of one provider which sought to bring together different “important actors” (such as schools, integration services, families) and the individual to draw out “the unconscious values” of each party. This was seen to create “a space to bring different identities together” and to help each group to “understand” and become “acquainted” with other values and perspectives. It was also described as facilitating trust and enabling “an adaptation process” that allows for “home cultures” to co-exist in the settlement space as people adapt to “living in a Western society”.

The importance of family and extended family perspectives in certain cultures, was a further consideration. There was “a need”, some said, to bring parents into consultations but to do this in a “sensitive” way; “you have to put it to the [family], you have to ask which person can begin or which one can say how they feel, or what is the problem”. It required an overt “welcome” of this input, “acknowledging respect and thanks for their contribution…”. One service explained that they did this even where parents might remain, in their home country,

> *“We cross-*borders*… so that we make that connection in terms of you know, how do the parents see what’s important in educating their son… so we can actually learn from that and then we can have more bi-cultural identity…”*.

In describing how they broach this contact, they explained,

> *“We don’t say, ‘well listen, we are psychologists’ but it’s more like, you know, ‘Your son has arrived [here]. He’s residing here’ so the parents themselves are very grateful, extremely grateful that they know like, okay, this person is actually asking me about my son and about his wellbeing… so just the fact that you are talking as person to person, you easily get into, yeh, a very close relationship…”*

Interpreters too were acknowledged as contributing to the intercultural exchange. One service talked about efforts to work with people that understood their therapeutic frame. There was the sharing of case files, “so that I can prepare, research and ensure that I [interpreter] have the full terminology of the specialist clinical language that will be necessary”, conversations with the psychologist before and after a session, “because heavy stuff is happening, and we [psychologists] are trained in not taking it in, but interpreters not”.

An interpreter who worked in this way described the approach as a,

> *“Protection that the psychologist gives to the translator… it’s a way to negotiate what happened and what was in the session. It’s a way to protect the translator from the affection that he gets in the session. It’s a very sensitive thing as a translator for these patients, psychologists’ patients, it’s very important to keep a space between you and the patient. The story of it. Try not to get involved in it…”*

There was the attempt to “fix” an interpreter to a particular client in order to build a “stronger” therapeutic environment creating a context in which the interpreter could be seen as a mediator of therapy, bringing a skill and cultural sensitivity to the therapeutic encounter; “each time we sit together… and this person, this patient he will feel more and more comfortable to talk and say everything inside him. Because he trusts the translator as he trusts the psychologist”.

Interpreters talked about the responsibility they felt to maintain trust and bring cultural understanding to an encounter. One interpreter used the example of gynaecology and the use of the word, “vagina” where a female patient and her husband may both be present, “so this woman comes from another land that maybe they don’t accept this word. And especially [with] the husband beside her…”. The interpreter explained that if they were to translate directly what had been asked, it would be seen as “a big shame… it will be the last word that I translate and they will leave and never come back. So I have to say it in another way… that is more respectable to this person, more acceptable”. Shifting terminology, they made clear, means conveying this clearly to the health professional, “I directly re translate… and tell the psychologists what I have translated for her. What words I have used… everything must be clear.”

#### Individuals and their cultural sphere

Facilitating the connection of people with shared cultures, faith, experiences, and language were felt to enable “better” care or support an environment in which there was a greater balance of empathy and understanding. This was seen through bringing clients or patients together in group-based practices, including therapy, health education and social activities, “it really helped them because they had the same feeling, they had the same pain for their families and when they were talking with each other… when they shared these feelings… it helped them to feel that they are not alone”.

It was also seen through the engagement of peer or lived experience advocates, usually volunteers, supporting clinical workers or group activities, providing interpretation, and supporting service and local navigation. This was seen to create a more “welcoming” environment, in which individuals could feel “safe” and “understood”. When someone “with a shared experience says, ‘I know how you feel…’ it’s because genuinely they do… I think it does make a massive difference… and can also give hope”.

> *“The advantage of being hired on the lived experience is you empathise with the people and even on the difficult patients who are finding it difficult to speak out, or to mention, or to open up and say this is what I need, you can actually read them and say, look I am here because of one, two, three and to be honest I am still on the same journey that you are”*.

There were lessons learned from COVID-19 vaccine outreach work that had been delivered alongside “members of different communities that knew their communities”. Social prescribing supported by peer volunteers,

> *“So we’re working really closely with the social prescribing team who are looking at getting volunteers to help at hotel sessions whether that be making appointments or helping with immunisation sessions… so it’s looking at how we can utilise their skills as well and they can become advocates for explaining things but also that’s that trusted member of the community that can… you know… be a positive link.”*

Bi-cultural workers in one service were described as people with understanding of community development or social services, who had strong links within the community (“they’re a community leader”) and who were able to negotiate and bridge between “mainstream” and their particular communities. They were also described as being well placed to “identify gaps that may be able to be met in ways that [services] haven’t looked at”.

There was some reference to practice managers, receptionists, nurses, and therapists from shared or similar communities, being able to better help navigate and bridge understanding, and though no service described the intentional recruitment of staff with minoritised characteristics, this was reflected across staff more broadly in many of the services. Gender however was acknowledged as a factor in certain decisions: a male coordinator was employed to help navigate some of the more complex contexts, such as family planning and faith perspectives; the benefit of flexibility to specify the desired gender of an interpreter, especially for women’s health was described; and one worker talked about being able to call on her (male) colleague in certain situations,

> *“So many times I’ll ring him when I’m in the hotel… I’m like, ‘do you have a moment, I’ve got this patient here’*. He’s *Arabic. Even if he’s not Arabic, he’s male, can I task you to be helping out with this, so we’re working hand in hand like that.”*

Though one worker pointed out, “it’s not the experience of the whole population” the propensity for someone with a shared background and language to “know what it means… to have the experience” and the understanding of “what’s going on in my society”, was widely recognised as a benefit to service users (and to services as a whole). In the delivery of mental health care particularly, this was articulated by workers from several services, with shared background/languages seen as “providing that assurance”, helping to reduce stigma and build trust in a service, “no, no, no you’re not nuts… and no I work here also” and that some activities, are just “better held with someone who knows more about the culture, knows more about the language”. One worker, who described themselves as coming from a “similar culture to the service users” reflected,

> *“I think it’s amazing that some of my colleagues have had the same lived experiences, because I remember when I was younger, I accessed psychotherapy but I could not relate to my counsellor at all, and I absolutely hated it. It was only when I went to uni and I thought ‘oh, okay, culture does make…’ and I’d just thought I hated the idea of psychotherapy because of my culture, but it wasn’t, it was more because I couldn’t relate to my counsellor… and I think in any role you need to be able to relate”*.

Working in “the mother language” of patients was seen as helping the therapeutic exchange to be more sensitive,

> *“In every language there are words, expressions, that we cannot translate. So it’s not easy for people sometimes to express themselves, say those words or those expressions, their thoughts, their feelings in other languages. We cannot translate it….”*

> *“It instils more confidence… it’s much more trustworthy because an interpreter communication has always those flaws, a risk of excluding important detail… Working with an interpreter is not the best solution. I mean it’s the best solution given the circumstances that we do not speak each other’s language but…”*.

#### The individual

There was some emphasis on being clear that “every migration tells a different story”, that people’s experiences and cultures should never be homogenised, and the value of making an effort to develop an understanding of each individual’s context. *Showing an interest* appeared instrumental to each service’s approach to care. That might be following trauma-informed principles or simply acknowledging someone’s “terrible” or “very difficult” situation. There was an interest in current events taking place in someone’s home country, though handled carefully, with several workers explaining that they don’t tend to go into much detail about “someone’s journey”. It was common to show an interest in what people used to do in their home country, to “ask about [someone’s] affinities and what [they] liked” and this was seen as a way of understanding the person and their cultural habitat. A “genuine curiosity” of others’ worlds, creating more equal relationships of “trust” and “reciprocity”, “in this way he is teaching me so we are the same… and this is going to create a better self-esteem”.

People talked about taking time to understand someone’s “rituals, traditions, behaviours, that come from their cultures”. In different contexts this seemed to be a clear priority, “just get to know them… what they want to do, what are their expectations… [and] what can be achieved together”. In this way, volunteer advocates talked about giving people,

> “*A lot of time… we stay and then chatting with them and making sure everything is okay… we talk about their problems… what makes you happy? They talk to us about finding places… we make sure you’re really happy before you leave this building. That’s our priority. We don’t work with minutes or with time…*”

This interest in the individual came with the practice of facilitating, supporting or signposting in response. Here social prescribing services were important: “It’s all about connecting you with your community”. It requires placing the individual in a space of autonomy, creating the conditions for “people themselves who have the wisdom to resolve their problems” to be “part of defining the solutions”.

A further rationale for community-based consultation, was the need for,

> *“Honouring the experience and the knowledge of the refugee themselves but it’s also kind of capacity building of the social assistant. Not to see a victim but to be culturally sensitive. To be aware of the migration experience. Not to start from I’ve got this man before me he’s a refugee.. but who was this man in his home country.. what can we continue. Because the mourning process is helped by continuation… So okay he was a lawyer, his diploma is not recognised so he cannot be a lawyer but there is some kind of analytic capacity so how can we translate it in our labour market”*.

### The individual and their circumstances: caring communities, coping and resiliency

The complexity of patients’ contexts was widely recognised and practice actively sought to engage with, influence, or support an individual to navigate their wider situation. There was acknowledgement to patients of the “many stress factors” shaping their experience,

> *“A lot of the symptoms that we think could be from a psychological origin [are] much more related to the mourning process of migration… it’s a loss experience and the stress of acculturation… you should not come to a psychologist; in that sense, a teacher or social assistant is much better placed to resolve this stress”*.

The same interviewee went on to explain,

> *“Talking about wanting to have your job back doesn’t help. It’s much more helpful to have a good social assistant thinking about what you are able to do, and to translate what the refugees still can do, into the new environment. ‘Okay, this is possible… this is not possible…’”*.

People talked about clients as “disconnected… from all the resolving or the problem-solving skills… problem-solving resources” of home and that it was these that services and receiving societies “have to put in place through informal or formal aid so that [people] can at least, get back on their feet and solve their problems themselves.” There was a commonly shared view that “it takes a society to heal war trauma” and an emphasis upon “prioritising daily stressors”. This included giving attention to “environmental factors”, “basic needs”, a “good lawyer”, “schooling”, “leisure activities”, “financial situations”, “barriers of language”, and “enforcing social contact”. These were seen as “instrumental in improving people’s circumstances” and “for stabilisation… coping mechanisms… as long as you’re in the asylum procedure”. This was often given as justification for wellbeing aspects of provision or work that took place with other sectors to support redress of these conditions. One interviewee described the need to affirm the importance of reconnecting people with their inner resilience,

> *“First of all, is that you know, people have already come from a very difficult area and sometimes… because they have been through so much, they don’t see their own strength anymore. So that’s where I try to have the person relate themselves back with how they have overcome already so many obstacles and that they have it in them to make something good out of this bad.”*

Services actively created the conditions for social connections such as through group activities, the involvement of peer advocates, and social prescribing. Making possible the connections with communities, faiths, interests, nature, sports, language classes, learning, training, volunteering and work opportunities were all important, supporting the capacity, through knowledge and confidence, for people to independently engage with services and make more informed decisions about their health: “[People] just need a network around them to support them in adjusting into a culturally new environment”, “when [these] things start to improve their mood starts to improve…”.

A “systemic view” of health and of the further deterioration of mental health explained services’ efforts to engage frontline workers in schools, social welfare and housing. Making those responders, who were seen as having a more confidential relationship with individuals, “aware” to consider “basic needs”, “resiliency”, and “how trauma works, and this is what you can do” was part of a multi-layered perspective and focus on “changing the rules and advocacy”. Each service in some form drove broader shifts in local responses: meetings with dental hospitals and community mental health teams “to explain the needs of our clients”; highlighting referral procedures that don’t recognise particular “vulnerabilities”; advocacy with housing, “to get a family with children with disabilities living in a hotel room into a proper house”; “myth busting” and increasing understanding of pre-arrival experiences; reminding colleagues that interpreter services “are free, here’s how you use it”.

A free Helpdesk for care providers offered by one service was available for outside professionals to seek advice and new perspectives on a specific case. Another service offered MDT support to other areas, and there were offers of formal and informal training, intervision and supervision to external agencies or individuals.

### Tensions in practice

#### A systems battle

In constant tension with services driving a *community of care* and a permanent point of bordering for individuals was what we have termed, a systems battle; immigration policies and institutional structures governing the broader care and social adjustment of patients and clients and diminishing and frustrating impact. At a functional level, “demystifying” the “terribly entrenched bureaucratic structures” of health and societal systems and supporting service navigation was a central activity provided by most services.

Despite variabilities for people with different immigration statuses or nationalities and across the different national contexts studied, racialised discrimination and xenophobia in health systems, community aggression, violence, including in schools and on the peripheries of accommodation sites were all acknowledged by service users and workers. But it was the hotel regime, that “adds to people’s stress” that was of greatest concern.

One person told us, “it’s really, really helpful to just get out” get involved in things and “see other places” but accommodation was often isolated, confined to an A-road and supermarket only or an unaffordable bus ride to and from services and communities. Faith appeared to be a strength for some people, as were volunteering opportunities. Some talked about “deep depression” … “you’re in the same room 24/7” and workers talked about witnessing a “loss of hope”. People smoke in shared rooms, sometimes of up to nine people, often with no windows or “sometimes they will open a crack”. People tell us that in some rooms where there are windows, there is often no way to block out the light. People described “no opportunities or activities inside the hotel… nothing to be busy with… no single opportunity just I can run on the main road up and down”. Everyone talked about food, including food affecting people’s mental health. This included the sense of a “loss of role from what they were used to… looking after the house, cooking their food…” and the sense of dehumanisation. From daily meals served on throwaway plates with throwaway cutlery and throwaway cups. People explained how there is often “no food from 5pm to 9am”, that meals are only ever “chicken and bread”, and some resorted to “living off biscuits” because there is no halal information, and people don’t trust the food nor what they are told about it.

> “*The diet that [people] would have… they’ve lost… you’re eating food that you’re not even used to… that probably you think is making you unwell. Like someone’s diabetes and eating chips every day is not going to be helping anyone*”.

People talked about coughing… Flu… COVID symptoms but no COVID tests. You cannot sleep either, several people tell us because of your own nightmares or because of someone else’s. One worker offers some techniques for managing nightmares but later explains, “they need at least a proper environment… in a hotel, therapies won’t work… these are very, very hostile environments”.

There was often talk of people living “in constant fear that somebody’s going to knock on the door and say, ‘get your things together, there’s a taxi outside’”. People are moved without being informed in their language what is going on, often with no warning and other times *with* warning but then it is not carried out. Workers talked about the impact that this has on people, “on their levels of anxiety”, “on their health care”, away from “the trusted service”, “split up from family”, they will “lose their friends”, “the networks of support” that people have built up, they will “miss out on their health assessments, or their care is disrupted….”. One service explained, “you know, they’re all human beings they’re not just numbers… what we’re sort of doing is re-traumatising them through the system… we’re disempowering them”. One service had developed a handheld health record as a means to address some of the disruptive consequences of patient relocation meaning that the physical record would remain with the person to facilitate continuity of care once relocated.

These spaces are *policed* by men in high-vis jackets, lots of them, steel-toed boots, walkie talkies; some wear stab vests. You get mixed impressions as to *whose* safety they might be protecting. In one hotel the toilet is out of service, with some suspicion that this is rather a ploy for preventing people congregating in the only shared space, the vestibule of the hotel; in most hotels there is nowhere to congregate at all. In another people tell us that only recently have rooms and showers started to be cleaned, people tell us it is a balance between challenging and retaining a relationship with these systems.

More than health care, what people really wanted was “to work… to have my papers as soon as possible…” *If we can’t change that… you’re shaking your head…*“Nothing… nothing actually… nothing, it just depends on yourself and how much you can handle…”.

> *“You’ll not starve, you’ll not sleep in the street but you’re not allowed to work… you’re not allowed to do anything… as an adult, that’s devasting. No services, no* activities*, no nothing, just stay in the hotel until [they] decide. Well, that’s okay if it’s a few months but sometimes it drags to years. It drags to years and then you are mentally demolished, and you feel that you are worthless”*.

> *“I think another person who doesn’t understand how it is to be sitting there. Everyday you’re looking at your phone. Every time an email comes in, you’re checking it’s the Home* Office*. When you miss a call, you get frustrated, what if it’s the Home Office… That gets to you without you knowing it”*

Workers talked of “sometimes… feel[ing] like you’re flying by the seat of your pants and putting out spot fires”, that they are merely keeping a patient’s “head above water”. Several questioned “the cost”, “the work”, and “the burden” on health services for “keeping people like this”.

#### Systemic cohesion and fracture

Partnerships and relationships were a “vital piece” of each model of care; between (re)settlement services, NGOs and community agencies, schools, social workers, carers of unaccompanied children, and some accommodation sites. GPs were sometimes integrated or strongly associated with services and usually co-located, providing mostly informal, supportive relationships both ways, clinical support and advice, the shared or co-informing on patients’ initial health assessments, and receptive to concerns.

> *“We are able to phone practice managers, discuss a particular case or a general issue with [a] local practice, prioritise a case of concern, and they will listen, they will take us at our word, you know”*.

A programme of refugee health fellows, a rotating group of on-call specialists (such as paediatricians and infectious disease specialists) was associated with one service and provided support and secondary consultations and were seen to “unblock the pathway” where there was concern about urgency or a particularly vulnerable case, “sometimes you can call the fellow and they’ll wander down to the emergency department, ‘I’m sending them to the ED now but can somebody when they get there, just like, fish them out and make sure they get where they need to go because they’re really ill…’”

Formal multidisciplinary meetings enabled broad issues “to be discussed”, plans “formulated” and “plugged into the local networks”. Some services talked of stakeholder meetings for “organisations that are interested in refugee health” some well attended, others explained that “health [and] accommodation services … sometimes just fall off the radar”.

On the whole, good relationships appeared to have either been long-existing, “like who do you know…”, or had relied on working very hard,

> *“It’s very much on their terms whether we have that good relationship… [we] try to change things for the positive and create systems change more than to criticise or condemn… obviously we do at times, we have to make complaints and raise quite serious issues but on the whole we bite our lips and try to do what we can to improve especially with the NHS as well… it’s about improving the system rather than condemning them for what they’ve done wrong”*.

There was some indication of benefits where services could embed staff or build strong connections with regional/state/national structures. Some strategic “influence” with access to decision makers, commissioners, taking that “voice [from] on the ground”, trusted across the different sectors and institutions and in a position to “nudge” decisions, funding, and practice.

There was a feeling that good things happened because of individuals and not by design and concerns were commonly raised relating to the coordination and communication of information, responsibilities and commitments across broader systems and services. Data was seen as a factor here,

> *“I think that these individuals get lost in data sets… they’re not coded as asylum seekers for the most part, so there’s no way of identifying how many asylum seekers are registered with GPs… And because they are one patient or a small number of patients in a list size of thousands, they don’t get noticed, even within the practice. And because they’re looked after by generalists, not specialists, again they don’t necessarily understand the needs to identify it or advocate for their needs. So, I think all of those things mean that they’re quite invisible”*.

There was talk among service users and workers of an abdication of responsibility, particularly within accommodation settings. Left often to other residents to care and look out for their peers. Who will call the ambulance? Who will check on each other? We hear about a young person,

> *“He described a terrible journey… ‘I am 17 years. The Home Office decided to add 10 more years on my age. I’m now 27 and I want to kill myself’….Do we call for adults or do we call for the children… Adult services… Crisis Help… CAMHS?”*

The young person ends up at A&E,

> *“If he’s a child he’s not supposed to be in a hotel, he’s a minor…”*

Over the phone, Crisis Help tell them,

> *“‘Please make sure there are no sharp objects in his room… a health hazard’. We can’t do that. We don’t have the authority to do that. We ask the hotel to do that, the hotel team are like, ‘we can’t we’re not allowed to do that’. We ask the housing team as well, they’re like, ‘We just accommodate them, we cannot go and do a search in their rooms’. So, it’s a complicated issue. You know. We’re standing there and we know he’s at risk but how do we help.”*

Suicidal thoughts and self-harm are commonly discussed. Who strategically is thinking about refugee patients and the health care of forced migrants, “there’s nobody highlighting them and they’re lost in the whole picture… commissioners really don’t have any idea”.

## Article summary and discussion

This qualitative analysis draws on five wide-ranging, transnational case examples. It sheds light on a range of mechanisms of particular relevance to regional and local commissioners, service leads, caring practitioners and civil society groups, that can be enacted, despite often constricted and hostile contexts and settings, to support the health and health care experiences of forced migrants.

We have suggested that these mechanisms coalesce around a set of shared *attitudes of care* through which *innovating & adapting; flexing for patient need;*and creating *spaces of trust* thread through the design and operational delivery of health care activities.

We shed light on the considered flexibility with which services can operate; flexing as a component of active learning, responsive to the changing service user and their individual needs, and operating in immigration environments in constant political flux. Our analysis shows that this capacity for flexibility stemmed from ambitious organisational attitudes of innovation and support. There was a freedom for reflexivity in how the everyday interactions of care operated and dedicated organisational practices of learning and rethinking offers of care. There was a fluidity in workers professionalisms, they were able to bring in ideas and have autonomy to explore the best responses to the people in their care, and there was some active resistance to ethnocentrism and the conditioning inherent in professional training.

This commitment to self-reflection aligns with broader health inequity scholarship which increasingly encourages a “self-reflection in personal biases, privileges, and power imbalances in interaction with patients”^64^ and has begun to acknowledge the structural systems that have long framed and disprivileged the social contexts and institutional interactions for racially, ethnically, and other minoritised societal groups. To move this forward, Abubakar ^65^ suggests we need, as well as legislation and race equity policies, a new set of activities through which “individual, organisational, and community change, and movement-building” can take place. What our research might contribute here is an articulation as to how some of these activities could be enacted; we have shown possibilities for divergent approaches to patient care, even in public sector contracts and contexts, and that this can largely be done through enabling and engendering reflexivity and flexibility in health care practice.

At a very basic level, the need to counter broader fear and unease in the displacement and immigration context, to actively create spaces of trust and safety through reassurance, transparency, avoiding known barriers, valuing the patient perspective and having patience and time, all appeared vital mechanisms in the work we studied. First encounters mattered and as others too have documented,^8,66,67^ a *real welcome*, taking different forms through reception spaces, a “welcoming smile”, peer support, an *Arabic greeting*, a *Farsi welcome*, a cup of tea, a glass of water, should not be underestimated.

A commitment to making communication work (high use of in-person interpreters, full utility of interpreters on the telephone, valuing interpreters as a vital component of the intercultural exchange that can bring trust and sensitivity to a clinical encounter) concurs with wider literature on the importance of dominant language proficiency as both a post migration stressor,^68^ a concern of clinical risk and medical error^69–71^ and multilingualism as central to health equity, self-expression and equal participation.^72^ The study has drawn attention to the benefit of a bilingual and bicultural workforce and the less-understood role of non-specialist bilingual peers in health care contexts, the latter, part of a broader informal offer of care and support; on reception, in waiting rooms, co-delivering care, and through social prescribing. The delivery of clinical care in the mother tongue, especially in the therapeutic exchange was also highlighted, the sensitivities of expression and feelings not to be underestimated in their importance. More broadly, other bilingual and/or bicultural workers offer a critical contribution bridging gaps between cultures and the health service and the other way, negotiating new forms of operation, decisions framed with better understanding, new perspectives on where the gaps or disconnects exist, and other ways of doing things. This largely echoes what Burgess et al^73^ highlight in their case for methods of bi-directional co-production, creating “welcoming and positive” statutory spaces that are “safe to enter”.

Maintaining connection with native identity and traditional values has been well documented in the context of migration,^74–76^ particularly forced migration where loss and mourning are supported by the maintenance of shared connections.^45^ Similarly, we identified that by locating care within a persons’ *cultural sphere* (group activities, peer support, bi-cultural workers) this could create dynamic spaces of shared experience and community that were felt to nurture understanding and hope, aide the negotiation of new connections and provide a bridge with a particular locale, including with the services studied and other health care providers. These alliances however should be determined by the individual themselves and as Fennig and Denov^21^ identify (in the context of working with interpreters from a patient’s community), with a particular sensitivity to potential hierarchical social norms and political, religious and ethnic factors, which may create for example, fear of moral judgement or increased anxiety around confidentiality. Nevertheless, our findings support wider observations that there may be value in exploring the integration of different (informal and formal) strategies and sectors into the design of health systems, particularly as means to address the health concerns and the structural inequalities and inequities impacting superdiverse populations.^77^

We also identified a pattern of *intercultural exchange*, conscious practices within micro/meso interactions that *give* on the part of *receiving* communities and created space for a negotiation for how diversity in identities, perspectives and understanding could co-align or co-integrate. Both play to the responsibility of health care providers to see a role in supporting the dynamic process of integration,^78,79^ enacting what Phillimore^44^ discusses as “refugee-integration-opportunity-structures”. This was seen in a commitment on the part of providers to ensure that people know how *this* society functions, its (health) systems, norms, and expectations. It was also shown in how care providers actively welcomed new understanding themselves, through a humility and capacity for inquisitiveness – prepared to become aware, sensitive and knowledgeable about others: *actively* creating spaces for exchanging cultural understandings; showing interest; welcoming the contribution of interpreters, peers and bilingual and bicultural colleagues (as discussed above); and through self-reflection.

We point also to an essential reflexiveness in respect of *individuals and their circumstances.*As others^44,80^ have discussed in the immigration context, this requires heightened recognition of the relationship between someone’s ill-health and the social structural dynamics and factors in which they are located. This recognition, as well as efforts to facilitate coping, resiliency and advocacy for change was often indivisible from more clinical aspects of the care that we studied. Again, the research highlighted that an inquisitiveness to people’s individual contexts and their own understandings of solutions, a prioritisation of what can be done about daily stressors, basic social, cultural and future orientated needs showed that there is a capacity for this broader interest to be built into traditional health care responses. Though advocacy in health care by which clinical professionals see a role in the broader context of their patients is fairly underexplored,^81^ we have shown that multiplicity in professional practice is possible; whether this be a nurse leaning towards social aspects of care while also ensuring efficiency in clinical prescribing, a psychologist supporting wider colleagues and collaborators to consider structural and social factors and good culturally sensitive interactions, or a GP engaging with housing services. Practitioners had a visible eye on the best collaborators in a patients care, having to-hand the relationships for a societal response; good intersectoral relations particularly with the community sector, social workers, schools, settlement services and clinical specialists arguably facilitated by the capacities of flexibility influencing practice and driving a natural divergence both within and across fields of care. Others have indicated that institutional arrangements at the local and country level matter for the establishment of such collaborations but that this requires the integration policy field and local policy forums to include a diversity of actors including immigrant advocacy bodies as respected partners in order to drive collaboration and successfully generate shared perspectives and projects.^82^

Finally, though the aim of the study was not to interrogate further barriers to care, our analysis highlights the tensions around which most services were required to navigate and compensate in order to maintain their handle on providing or supporting access to care. We articulate this in part as a *systems battle* to which much of what services were required to do revolved around compensating for both exclusionary (policy) strategies and what Norman^83^ terms “strategic indifference”. Though to differing degrees, hostile immigration environments were operating across the national contexts and this was seen to disempower people in states of precarity materially and psychologically and hinder their social inclusion, including health inclusion. In such processes, Norman suggests that there is a rational and strategic inaction on the part of the state and a reliance on others (usually non-state actors) to step in and provide what governments do not explicitly deny but make difficult to achieve.^83^

The flexing of services in their navigation of this has some overlap with what Tomkow^84^ and others identify as ‘tinkering’; a practice whereby health care workers in often (economically and ideologically) resource-poor and unpredictable settings use workarounds or make-do in order to get the job done. Though we would maintain that the general trend to flexing care in the services we studied was centred around those reflexive and learning practices that operated not *because of* systems but as an important capability *within their* systems, we suggest that these practices need to be clearly defined and should be distinguished from other flexibilities which ultimately tolerated broader structural inadequacies (bloods taken, for example, in an accommodation staff room with security sleeping nearby and care in a corner of a canteen). So while services often acknowledged that spaces and systems were not conducive to appropriate care, despite advocacy and despite negotiation, there was in effect at times an impasse between services and external agents (government departments and contractors) which ultimately enforced a tolerance, without which, those we interviewed believed there would simply be no care at all. This precarity does leave us with important questions around strategic inaction and the risk, as Tomkow points out, of making-do and further embedding discriminatory or sub-standard practice as usual care.^84^

Ultimately, this seems an appropriate reflection on state attitudes to immigration policy, not only but particularly in the UK; extensive waits in processing asylum claims, a contingency accommodation regime, and isolating and restrictive policies and practices and our evidence certainly echoes what several recent reports have articulated around risk of harm produced by conditions in these spaces and contexts.^34,85–87^ We would argue that this places unacceptable limits on the achievement of health and fosters wasted resource within health care systems and suggest this justifies a rapid review of practices and their implications on short, medium and long term outcomes both for individuals and health providers.

## Limitations and future directions

We see our stakeholder engagement, our lived experience contributors and multisector team as a particular strength of the project and attempted to ensure that this involvement was meaningful in both directions. Our case examples engaged critical perceptions from across a range of international jurisdictions and contexts and we highlight this as a further strength and suggest that transnational strategies for learning and the sharing of good practice should be embedded in practice and policy development.

Though we conducted more than 50 hours of interviews and engaged in five days of observations and informal conversation, observations were brief which we acknowledge has methodological implications. Our direct engagement with service users was lower than anticipated and only with those from some services. This resulted from unexpected restrictions including those imposed by accommodation providers where services were delivered, concern for burden on patient groups and service pressures). Though we also interviewed workers who were former service users, we suggest further qualitative research should prioritise evaluative methodologies that are able to consider, in more depth, the experiences and health care impacts of similar services, particularly as they relate to the medium and longer term. We note also that some interviews relied on interpreters and note taking. In our documenting, through the process of translation and in our validating of the data, we remained conscious of the risk of interpretations altering respondents’ meaning as we moved between word transition and linguistic structures.

We provided opportunity for included services to validate interpretations and correct misunderstandings, a strategy to support trustworthiness in qualitative research.^60,61^

For refugee communities, we suggest that there is further exploration of the role and training of non-specialist peers in health care practice in a range of facilitative and delivery roles, including interpretation. For practitioners, we suggest that research is conducted to better understand (1) how we motivate, engage and leverage alliance-building relationships across a broad constituency of caring practitioners and (2) the models and possibilities for embedding intercultural exchange, competencies and reflexivities in both professional education and professional practice.

## Conclusions

There is a primary need for reflexivity in health care practice and a radical commitment to intercultural exchange, humility, building trust and good communication. Environments that enable good health and enable people to live lives of meaning are also vital and integral to a health care response. These factors are possible through micro-flexibility with patients and systems flexibility which sees the opening-up of health care practice to include a broader range of caring practitioners that can, and should, through intentional and interconnected practice, contribute to the health care of forced migrants.

Future work should focus on the health and health service implications of immigration practices; the inclusion of peers in a range of health care roles; alliance-building across unlikely collaborators and the embedding of intercultural exchange in practice. Findings of this study are supported by our systematic review (publication forthcoming).

## Contributors

**Amy Robinson** Conceptualisation; Funding acquisition; Project administration; Methodology; Workshops & events; Investigation; Data curation; Formal analysis; Visualisation; Writing – original draft. AR acted as guarantor.

**Ziaur Rahman A. Khan** Project administration; Workshops & events; Investigation; Formal analysis; Writing – original draft.

**Kofi Broadhurst** Data curation; Formal analysis; Visualisation; Writing – original draft.

**Professor Laura Nellums** Conceptualisation; Funding acquisition; Project administration; Writing – review and editing; Project supervision.

**Gisela Renolds** Conceptualisation; Funding acquisition; Project administration; Workshops & events; Writing – review and editing.

**Bayan Payman** Conceptualisation; Project administration; Methodology; Workshops & events; Writing – review and editing.

**Professor Andrew Smith** Conceptualisation; Funding acquisition; Project supervision.

## Patient and public involvement

We actively attempted an inclusive project approach that attempted to soften the boundaries of research as an externalist practice. Our project team was multisector and included people with refugee backgrounds, refugee and asylum-seeker support services, health and other public service providers, and academics. This brought a diversity of perspective to decisions and practice; helped to emphasise the importance of clarity in communication which needed to be succinct and framed for different audiences; helped to clarify different expectations we might have of different participants and the lines of enquiry we should follow. We sought additional input from others with lived experience in relation to planning and co-facilitation of community workshops and planning content and reviewing of participant materials.

We delivered a range of community workshops, conversations and events with refugee communities (46 individuals with a refugee background were involved across 4 sessions) and the voluntary, public and private sector. These were intended as a means to help to support the validity and relevance of our research in both the needs of forced migrant communities and those of caring practitioners in the field and was an attempt to reach some of those who may be able to utilise our learning. A summary of these events is reported in a forthcoming report. Forced migrant community members who participated in the research or attended workshops were provided vouchers to acknowledge their contribution.

## Acknowledgements

We extend significant gratitude to participating services and individuals for their time and their willingness to share their experiences, stories and reflections. We want to thank Faten Almregawe and Aryan Kareem for bringing their lived experience perspectives to different aspects of the study, Dr Amy Lee and Antony Chuter for their support as Primary Care and patient and public involvement advisors, respectively, and Ashley Smith and Michael Pritchard for support with researching and cataloguing case examples. In addition, we would like to thank Sharon Lewis for support with funding acquisition, delivering workshops and community engagement.

## Data availability statement

Further data are available on request from the corresponding author.

## References

1. Clark RC, Mytton J. Estimating infectious disease in UK asylum seekers and refugees: a systematic review of prevalence studies. J Public Health 2007;29:420–8.

2. WHO. Report on the health of refugees and migrants in the WHO European Region. No PUBLIC HEALTH without REFUGEE and MIGRANT HEALTH. 2018. https://apps.who.int/iris/bitstream/handle/10665/311347/9789289053846-eng.pdf?sequence=1&isAllowed=y

3. Al Qadire M, Al-Shdayfat, N. Cancer awareness and barriers to seeking medical help among Syrian refugees in Jordan: a base-line study. J Cancer Educ 2019;34(1):19–25.

4. Drury J, Williams, R. Children and young people who are refugees, internally displaced persons or survivors or perpetrators of war, mass violence and terrorism. Curr Opin Psychiatry. 2012;25(4):277–284. doi:10.1097/YCO.0b013e328353eea6

5. Fazel M, Wheeler J, Danesh J. Prevalence of serious mental disorder in 7,000 refugees resettled in Western countries: A systematic review. The Lancet. 2005;365:1309–1314.

6. Aspinall P. Hidden needs. Identifying key vulnerable groups in data collections: vulnerable Migrants, Gypsies and Travellers, Homeless People, and sex workers. 2014. https://assets.publishing.service.gov.uk/government/uploads/system/uploads/attachment_data/file/287805/vulnerable_groups_data_collections.pdf

7. Kang C, Tomkow L, Farrington R. Access to primary health care for asylum seekers and refugees: a qualitative study of service user experiences in the UK. Br J Gen Pract. 2019;69(685):537–545. doi:10.3399/bjgp19X701309

8. Bradby H, Lindenmeyer A, Phillimore J, Padilla B, Brand T. ‘If there were doctors who could understand our problems, I would already be better’: dissatisfactory health care and marginalisation in superdiverse neighbourhoods. Sociol Health Illn. 2020; 42:739–757. doi:10.1111/1467-9566.13061

9. Cheng IH, Drillich A, Schattner P. Refugee experiences of general practice in countries of resettlement: a literature review. Br J Gen Pract. 2015;doi:doi.org/10.3399/bjgp15X683977.

10. Nellums L, Rustage K, Hargreaves S, Friedland J, Miller A, Hiam L. Access to healthcare for people seeking and refused asylum in Great Britain: a review of evidence. 2018.

11. Mann JM, Gostin L, Gruskin S, Brennan T, Lazzarini Z, Fineberg HV. Health and Human Rights. Health and human Rights. 1994;1:6–23.

12. Lustig SL, Kia-Keating, M., Knight, W.G., Geltman, P., Ellis, H., Kinzie, J.D., Keane, T., Saxe, G.N. Review of child and adolescent refugee mental health. Journal of the American Academy of Child and Adolescent Psychiatry. 2004;43:24–36.

13. Morris MD, Popper, S.T., Rodwell, T.C., Brodine, S.K., Brouwer, K.C. Healthcare barriers of refugees post-resettlement. J Community Health. 2009;34(6)doi:10.1007/s10900-009-9175-3

14. Bauer AM, Alegría, M. Impact of patient language proficiency and interpreter service use on the quality of psychiatric care: a systematic review. Psychiatr Serv. 2010;61(8):765–73. doi:10.1176/ps.2010.61.8.765)

15. Gadeberg AK, Norredam, M. Urgent need for validated trauma and mental health screening tools for refugee children and youth. Eur Child Adolesc Psychiatry. 2016;25:929–931. doi:10.1007/s00787-016-0837-2

16. Robertshaw L, Dhesi S, Jones L. Challenges and facilitators for health professionals providing primary healthcare for refugees and asylum seekers in high-income countries: a systematic review and thematic synthesis of qualitative research. BMJ Open. 2017;7doi:10.1136/bmjopen-2017-015981

17. WHO. Promoting the health of refugees and migrants: FRAMEWORK OF PRIORITIES AND GUIDING PRINCIPLES TO PROMOTE THE HEALTH OF REFUGEES AND MIGRANTS. 2018. https://www.who.int/docs/default-source/documents/publications/promoting-health-of-refugees-migrants-framework-and-guiding-principles.pdf?sfvrsn=289d4ae6_1

18. Burnett A, Peel, M. Health needs of asylum seekers and refugees. BMJ. 2001;322(7285):544–547.

19. Papadopoulos I, Lees S, Lay M, Gebrehiwot A. Ethiopian refugees in the UK: migration, adaptation, and settlement experiences and their relevance to health. Ethn Health. 2004;9(1):55–73.

20. Patel A, Corbett, J. Registration refused: a study on access to GP registration in England.2017.

21. Fennig M, Denov, M. Interpreters working in mental health settings with refugees: An interdisciplinary scoping review. Am J Orthopsychiatry. 2021;91(1):50–65. doi:10.1037/ort0000518

22. Daniel M , Devine C, Gillespie R ea. Helping new refugees integrate into the UK: baseline data analysis from the survey of New Refugees.. 2010. https://www.gov.uk/government/uploads/system/uploads/attachment_data/file/116069/horr36-report.pdf

23. Harris MF, Telfer BL. The health needs of asylum seekers living in the community. Med J Aust. 2007;175(11–12):589–592.

24. Ballatt J, Campling P, Maloney C. Intelligent Kindness: Rehabilitating the welfare state. Cambridge University Press; 2020.

25. Montgomery E, Foldspang, A. Discrimination, mental problems and social adaptation in young refugees. European Journal of Public Health. 2008;Volume 18(2):156–161. doi:10.1093/eurpub/ckm073

26. Clancy M, Taylor J, Bradbury-Jones C, Phillimore J. A systematic review exploring palliative care for families who are forced migrants. Journal of Advanced Nursing. 2020;76(11):2872–2884.

27. Erasmus E. The use of street-level bureaucracy theory in health policy analysis in low- and middle-income countries: a meta-ethnographic synthesis. Health Policy and Planning. 2014;29(suppl_3):iii70–iii78. doi:10.1093/heapol/czu112

28. Kyriakidou M. Hierarchies of deservingness and the limits of hospitality in the ‘refugee crisis’. *Media*, Culture & Society. 2021;43(1):133–149. doi:10.1177/0163443720960928

29. Weller SJ, Crosby LJ, Turnbull ER, et al. The negative health effects of hostile environment policies on migrants: A cross-sectional service evaluation of humanitarian healthcare provision in the UK. Wellcome Open Res. 2019;22(4)doi:doi: 10.12688/wellcomeopenres.15358.1.

30. DOTW. Deterrence, delay and distress: the impact of charging in NHS hospitals on migrants in vulnerable circumstances. 2017. Accessed 1 October 2022. https://www.doctorsoftheworld.org.uk/wp-content/uploads/import-from-old-site/files/Research_brief_KCL_upfront_charging_research_2310.pdf

31. Qureshi A, Morris M, Mort L. ACCESS DENIED: THE HUMAN IMPACT OF THE HOSTILE ENVIRONMENT. 2020.

32. Mayblin L. Impoverishment and Asylum: Social Policy as Slow Violence. Routledge; 2020.

33. Selvarajah S, Maioli SC, Deivanayagam TA, et al. RACISM, XENOPHOBIA, DISCRIMINATION, AND HEALTH. 2022;400(10368):2109–2124.

34. RefugeeCouncil. LIVES ON HOLD Experiences of people living in hotel asylum accommodation. A follow-up report. 2022. https://www.refugeecouncil.org.uk/wp-content/uploads/2022/07/Lives-on-hold-research-report.-July-2022.pdf

35. Eisenbruch M. The mental health of refugee children and their cultural development. International Migration Review. 1988;22:282–300.

36. Davies AA, Blake C, Dhavan P. Social determinants and risk factors for non-communicable diseases (NCDs) in South Asian migrant populations in Europe. Asia Europe J. 2011;8(4):461–73.

37. Bemak F, Chung, R.C. Immigrants and refugees. In: Leong FTL, Nagayama Hall, G.C., McLoyd, V.C., et al., ed. APA Handbook of Multicultural Psychology, Vol 1: Theory and Research. American Psychological Association; 2014:503–517.

38. Vandemark LM. Promoting the sense of self, place, and belonging in displaced persons: the example of homelessness. . Arch Psychiatr Nurs 2007;21(5):241–8. doi:10.1016/j.apnu.2007.06.003. PMID: 17904481.

39. Serneels G, O’Driscoll JV, Imeraj L, Vanfraussen K, Lampo A. An Intervention Supporting the Mental Health of Children with a Refugee Background. Issues in Mental Health Nursing. 2017;38(4):327–336. doi:10.1080/01612840.2017.1285969

40. Schonfeld DJ, Demaria, T. Providing Psychosocial Support to Children and Families in the Aftermath of Disasters and Crises. Pediatrics. 2015;136(4):e1120–e1130. doi:10.1542/peds.2015-2861

41. UN. United Nations Resolution 70/1. Transforming our world: the 2030 agenda for sustainable development. New York: United Nations; 2015.

42. EHRC. Making sure people seeking and refused asylum can access healthcare: what needs to change? 2018. Accessed 10 March 2023. equalityhumanrights.com/sites/default/files/people-seeking-asylum-access-to-healthcare-what-needs-to-change.pdf

43. Fiddian-Qasmiyeh E, Loescher G, Long K, Sigona NE. The Oxford Handbook of Refugee and Forced Migration Studies. Oxford University Press; 2014.

44. Phillimore J. Refugee-integration-opportunity structures: shifting the focus from refugees to context. Journal of Refugee Studies. 2020;34(2):1946–1966. doi:10.1093/jrs/feaa012

45. O’Driscoll JV, Serneels, G., Imeraj, L. A file study of refugee children referred to specialized mental health care: from an individual diagnostic to an ecological perspective. Eur Child Adolesc Psychiatry. 2017;26(11):1331–1341. doi:10.1007/s00787-017-0981-3

46. Juárez SP, Honkaniemi H, Dunlavy AC, al e. Effects of non-health-targeted policies on migrant health: a systematic review and meta-analysis. Lancet Glob Health. 2019;7(4):420–435. doi:10.1016/S2214-109X(18)30560-6

47. Porter M, Haslam N. Predisplacement and postdisplacement factors associated with mental health of refugees and internally displaced persons. A meta-analysis. J Am Med Assoc. 2005;294:602–612.

48. Bronfenbrenner U. Toward an experimental ecology of human development. Am Psychol. 1977;32:513–531.

49. Windsong EA. Incorporating intersectionality into research design: an example using qualitative interviews. Int J Soc Res Method. 2018;21(2):135–147.

50. Bradby H, Thapar-Björkert S, Hamed S, Maina Ahlberg B. Undoing the unspeakable: researching racism in Swedish healthcare using a participatory process to build dialogue. Health Research Policy and Systems. 2019;17(43)

51. Crenshaw K. Mapping the Margins: Intersectionality, Identity Politics, and Violence against Women of Color. Stanford Law Review. 1991;43(6):1241–1299.

52. Morgan SJ, Macdonald LM, McKinlay EM, Gray BV. Case study observational research: a framework for conducting case study research where observation data are the focus. Qual Health Res. 2017;27(7):1060–8.

53. Service RW. Book Review: Corbin, J & Strauss, A (2008). Basics of Qualitative Research: Techniques and Procedures for Developing Grounded Theory (3rd ed.). Thousand Oaks, CA: Sage. Organizational Research Methods. 2009;12(3):614-617. doi:10.1177/1094428108324514

54. Russell J, Greenhalgh T, Boynton P, M R. Soft networks for bridging the gap between research and practice: illuminative evaluation of CHAIN. BMJ. 2004;15(328)doi:10.1136/bmj.328.7449.1174

55. Polanyi M. The tacit dimension. Anchor Day; 1962.

56. Saurman E. Improving access: modifying Penchansky and Thomas’s Theory of Access. Journal of Health Services Research & Policy. 2016;21(1):36–39. doi:10.1177/1355819615600001

57. J M. Qualitative research design: an interactive approach. Sage Publications; 2012.

58. Braun V, Clarke, V. Using thematic analysis in psychology. Qual Res Psychol. 2006;3(2):77–101.

59. Potter JW, M. Discourse and Social Psychology: Beyond Attitudes and Behaviour. Sage; 1987.

60. Lindheim T. Participant Validation: A Strategy to Strengthen the Trustworthiness of Your Study and Address Ethical Concerns. In: Espedal G, Jelstad Løvaas B, Sirris S, Wæraas A, eds. Researching Values: Methodological Approaches for Understanding Values Work in Organisations and Leadership. Springer International Publishing; 2022:225–239.

61. Merriam SB, Tisdell EJ. Qualitative research: A guide to design and implementation. 4th ed ed. Jossey-Bass; 2015.

62. Birt L, Scott S, Cavers D, Campbell C, Walter F. Member Checking: A Tool to Enhance Trustworthiness or Merely a Nod to Validation? Qualitative Health Research. 2016;26(13):1802–1811. doi:10.1177/1049732316654870

63. Mercer J. The challenges of insider research in educational institutions: wielding a double-edged sword and resolving delicate dilemmas. Oxford Review of Education. 2007/02/01 2007;33(1):1-17. doi:10.1080/03054980601094651

64. Lokugamage AU, Meredith, A. Women from ethnic minorities face endemic structural racism when seeking and accessing healthcare. 2020. Accessed 2 March 2023. https://blogs.bmj.com/bmj/2020/03/05/women-from-ethnic-minorities-face-endemic-structural-racism-when-seeking-and-accessing-healthcare/

65. Abubakar I, Gram, L., Lasoye, S., Achiume, E.T., et al. . Confronting the consequences of racism, xenophobia, and discrimination on health and health-care systems. Lancet. 2022;400(Racism, Xenophobia, Discrimination, and Health 4):2137–46.

66. Hamed S, Bradby H, Ahlberg BM, Thapar-Björkert S. Racism in healthcare: a scoping review. BMC Public Health. 2022;16(22)doi:10.1186/s12889-022-13122-y.

67. Humphris R, Bradby H, Padilla B, Phillimore J, Pemberton S, Samerski S. After encounters: revealing patients’ unseen work through their pathways to care”. *International Journal of Migration*, Health and Social Care. 2020;16(2):173–187. doi: 10.1108/IJMHSC-07-2019-0066

68. Kartal D, Alkemade N, Kiropoulos L. Trauma and Mental Health in Resettled Refugees: Mediating Effect of Host Language Acquisition on Posttraumatic Stress Disorder, Depressive and Anxiety Symptoms. Transcultural Psychiatry. 2019;56(1):3–23. doi:10.1177/1363461518789538

69. Weiss L, Gany F, Rosenfeld P, et al. Access to multilingual medication instructions at New York City pharmacies. J Urban Health. Nov 2007;84(6):742–54. doi:10.1007/s11524-007-9221-3

70. Bischoff A, Bovier PA, Rrustemi I, Gariazzo F, Eytan A, Loutan L. Language barriers between nurses and asylum seekers: their impact on symptom reporting and referral. Soc Sci Med. Aug 2003;57(3):503–12. doi:10.1016/s0277-9536(02)00376-3

71. Flores G, Laws MB, Mayo SJ, et al. Errors in Medical Interpretation and Their Potential Clinical Consequences in Pediatric Encounters. Pediatrics. 2003;111(1):6–14. doi:10.1542/peds.111.1.6

72. Parente V, White MJ. Equity Is Multilingual: A Call for Language Justice in Pediatric Hospital Medicine. Hospital Pediatrics. 2023;13(3):e51–e53. doi:10.1542/hpeds.2022-007077

73. Burgess RA, Choudary N. Time is on our side: operationalising ‘phase zero’ in coproduction of mental health services for marginalised and underserved populations in London. International Journal of Public Administration. 2021/07/04 2021;44(9):753–766. doi:10.1080/01900692.2021.1913748

74. Acar B, Acar İ H, Alhiraki OA, Fahham O, Erim Y, Acarturk C. The Role of Coping Strategies in Post-Traumatic Growth among Syrian Refugees: A Structural Equation Model. Int J Environ Res Public Health. Aug 21 2021;18(16)doi:10.3390/ijerph18168829

75. Dangmann C, Solberg Ø, Myhrene Steffenak AK, Høye S, Andersen PN. Syrian Refugee Youth Resettled in Norway: Mechanisms of Resilience Influencing Health-Related Quality of Life and Mental Distress. Front Public Health. 2021;9:711451. doi:10.3389/fpubh.2021.711451

76. Wicki B, Spiller TR, Schick M, et al. A network analysis of postmigration living difficulties in refugees and asylum seekers. Eur J Psychotraumatol. 2021;12(1):1975941. doi:10.1080/20008198.2021.1975941

77. Phillimore J, Bradby, H., Brand, T., Padilla, B., Pemberton, S. Exploring Welfare Bricolage in Eurpoe’s Superdiverse Neighbourhoods. Routledge; 2021.

78. Fichter JH. Sociology. Chicago University Press; 1957.

79. EuropeanCommission. The Common Basic Principles for Immigrant Integration Policy in the EU. 2004. Accessed 2 February 2023. https://ec.europa.eu/migrant-integration/library-document/common-basic-principles-immigrant-integration-policy-eu_en

80. Ager A, Strang A. Understanding Integration: A Conceptual Framework. Journal of Refugee Studies. 2008;21(2):166–191. doi:10.1093/jrs/fen016

81. Abbasinia M, Ahmadi, F., Kazemnejad, A. . Patient advocacy in nursing: A concept analysis. Nursing Ethics. 2020;27(1):141–151. doi:10.1177/0969733019832950

82. Schiller M, Martínez-Ariño J, Bolíbar M. A relational approach to local immigrant policy-making: collaboration with immigrant advocacy bodies in French and German cities. Ethnic and Racial Studies. 2020/09/01 2020;43(11):2041–2061. doi:10.1080/01419870.2020.1738524

83. Norman KP. Reluctant Reception: Refugees, Migration and Governance in the Middle East and North Africa. Cambridge University Press; 2020.

84. Tomkow L, Prager G, Drinkwater J, Morris RL, Farrington R. ‘That’s how we got around it’: a qualitative exploration of healthcare professionals’ experiences of care provision for asylum applicants’ with limited English proficiency in UK contingency accommodation. BMJ Open. 2023;13(11):e074824. doi:10.1136/bmjopen-2023-074824

85. Jones L, Phillimore J, Fu L, Hourani J, Lessard-Phillips L, Tatem B. “They just left me”: Asylum seekers, health, and access to healthcare in initial and contingency accommodation. 2022.

86. Action R. Hostile Accommodation: How the asylum housing system is cruel by design. 2023.

87. RCPSYCH. Protecting the mental health of people seeking sanctuary in the UK’s evolving legislative landscape. 2024. Accessed 15 November 2024. https://www.rcpsych.ac.uk/docs/default-source/improving-care/better-mh-policy/college-reports/college-report-cr242_protecting-the-mental-health-of-people-seeking-sanctuary.pdf?sfvrsn=5a6f0faa_25

