## Supplementary material for "Mechanisms and attitudes in responsive health care for forced migrant communities. A qualitative study of transnational practice"

### Case study 1.

The Health Access to Refugees Programme (HARP) is a community outreach programme delivered by the Refugee Council in West Yorkshire, UK.^265^ With a focus on overcoming barriers to health care for asylum seekers and refugees, HARP adapted its community-based model to respond to the sharp increase in use of temporary contingency hotel accommodation in the UK in 2021. Working across both urban and remote suburban sites in West Yorkshire their focus is primarily upon health, health-related issues, health advocacy and awareness-raising but describe, because of high levels of need, additional drop-in support relating to medical and dental appointments, medication, and basic needs.

Predominantly charity and foundation funded they have no formal responsibility for facilitating access to health provision though this is a common activity of the service. The primary focus of the service is health education support, such as health access workshops to improve understanding of the NHS and its services, supporting people to access provision in an appropriate way and at the appropriate time, and providing ESOL for health and condition-specific health awareness sessions. The HARP approach includes capacity building, proactively reaching out to health services and other organisations to raise awareness and provide training, for example, around understanding needs and entitlements, and reactively, responding to issues raised by clients or partner services.

HARP offer opportunities for peer volunteers to receive training and mentoring to provide advocacy support to their peers. This commonly includes translation and directly supporting physical access to medical appointments and the collection of prescriptions. Peer volunteer advocates also support the broader training and awareness provided by the service.

HARP is delivered by a small staff team with volunteers working in and across different sites. The service works predominantly with asylum seeking males, but also with families and women in women’s only accommodation.

| **WHAT** | NON-CLINICAL, OUTREACH ADVOCACY, EDUCATION AND SUPPORT TO ACCESS THE UK HEALTH CARE SYSTEM | | | | |
| --- | --- | --- | --- | --- | --- |
| **WITH** | ASYLUM SEEKERS AND REFUGEES; LOCAL PUBLIC SERVICE TEAMS, EDUCATIONAL INSTITUTIONS, COMMUNITY GROUPS | | | | |
| **WHERE** | INITIAL AND CONTINGENCY ACCOMMODATION (MOSTLY HOTELS), WEST YORKSHIRE, UK | | | | |
| **HARP**  Direct health access (HARP staff)  Drop-in and casework | |  | **VOLUNTEERS (Former clients)**  Peer support |  | **CAPACITY BUILDING**  Proactive and reactive – responding to need |
| **SUPPORT** telephone communication with GPs, midwives, hospitals, dentists, mental health/wellbeing services, and social care services; making, querying or chasing up appointments. | |  | **SUPPORT** awareness raising and capacity building sessions including drawing attention to current situations within hotels and barriers experienced by asylum seekers seeking care. |  | **AWARENESS RAISING** with NHS and other public service teams and community groups supporting understanding of needs, entitlements, experiences, and working with interpreters. Undertaken via talks, staff meetings, training sessions and conferences. Face-to-face and online. |
| **UNDERSTANDING** health service letters and texts about appointments, referrals, and test results. | |  | **ADVOCACY** with hotel staff bringing to attention Home Office, health and accommodation issues on behalf of hotel residents. |  | **GP TRAINEE AND MEDICAL STUDENT** talks on refugee and asylum seeker needs in partnership with local university and health trust. |
| **UNDERSTANDING** directions to clinics procedures and expectations around appointments and waiting times. | |  | **FACILITATE** health care appointments through in-person reminders. |  | **SCHOOL/COLLEGE STUDENT** information sessions and awareness for those interested in medical school. |
| **EDUCATION + AWARENESS** through specialised health workshops incl. men’s health, women’s health, mental health, and maternal health; health service navigation through organised sessions and one-to-one engagement. | |  | **ACCOMPANYING** to medical appointments and pharmacy helping with locating, physically accessing, moral support. |  | **ADVOCACY** approach responding to issues flagged by clients and other services and raising concerns, including supporting the wider understanding of clients’ needs. |
| **ADVOCACY** with health services drawing attention to urgent medical and dental care needs; challenging charging. | |  | **ADVOCACY** to all services (including HARP) with benefit of language, cultural, faith and home-country understanding. |  | **STAFFING**  **Small team** moving across different accommodation sites supported with monthly clinical supervision. |
| **ESOL FOR HEALTH** encouraging learning of words that can help with communicating health issues and needs. | |  | **SUPPORT** understanding Home Office communications, asylum processes, removals, and transfers. |  |  |
| **SIGNPOST** to community groups and activity providers, such as climbing walls, Wildlife Trusts, gyms, kick boxing. | |  | **INTERPRETATION** Formal and informal. |  |  |
| **WELCOMING** new arrivals and provision of resources such as mobile phones, bikes and shoes (via donations). | |  |  |  |  |

HARP case example

### Case study 2.

RESPOND is a community-based holistic health screening and care planning service working with people seeking asylum across five London boroughs in the UK. Core screening and health planning sits within a flexible delivery model that works across a number of GP practices and contingency/temporary accommodation sites at any given time. Clients are actively approached by the team, usually by telephone and with telephone interpreters (where appropriate) and invited to an appointment. Text messages, verbal reminders within accommodation settings, and sometimes additional phone calls will be made as appointment reminders; flexibility and understanding are seen as a priority in supporting attendance.

The service team includes a health care assistant, Infectious and Inclusion Health Practitioners (with advanced nurse practitioner backgrounds), consultant leads in paediatrics and infectious diseases including specialism in safeguarding and looked after children, a matron providing management support, a GP also involved in research and service evaluation, and administrative, operational and service management support.

RESPOND take a trauma-informed and family-based approach. An expert MDT meets regularly, often involves wider community partners such as family support workers, social care, school nursing, health visiting, education, and the voluntary and community sector. The meetings offer opportunities for case discussion, clinical support, fast-track referrals to specialists outside of standard care pathways, and awareness raising (that might relate to COVID-19 outbreaks, a new resource, or an influx of new arrivals from a particular country). A virtual extension of this MDT service has recently become available to all services and locations across the UK.

Following screening, an electronic and handheld health plan including signposting and referrals, aims to support engagement with other services and address some of the health impacts of disruptions to care from the common relocation of asylum-seekers to different parts of the UK.

Integral to the core screening and care planning provided, the service relies on its capacity to flex and adapt to moving in and out of new locations, working in different ways and in different sorts of spaces, to continually engage and liaise with other services to identify new accommodation sites and appropriate GP practices, as well as consider new ways of engaging clients.

| **WHAT** | **COMMUNITY-BASED HOLISTIC HEALTH ASSESSMENT AND CARE PLANNING** | | | | |
| --- | --- | --- | --- | --- | --- |
| **WITH** | ASYLUM SEEKING ADULTS, FAMILIES AND CHILDREN | | | | |
| **WHERE** | INITIAL AND CONTINGENCY ACCOMMODATION SITES, PRIMARY CARE SITES, FIVE LONDON BOROUGHS, UK. | | | | |
| **RESPOND MODEL, UCLH** | |  | **ENGAGING CLIENTS** |  | **PARTNERSHIPS** |
| **Holistic health assessment and care planning** with asylum seekers within temporary accommodation. Includes a focus on physical health, infectious diseases, emotional and psychological wellbeing, sexual and reproductive health, child development and family functioning, access to play, education, and social support services or resources, trauma and safeguarding, oral and dental health.  Each area of focus will consider an associated response or action including multi-sector referrals (often rapid and outside of standard care pathway), booking and facilitation of access to other services, and some provision of resources and signposting. | |  | **Proactive ‘referrals’** with administrative staff constantly working to identify and connect with new contingency accommodation (hotel) sites. Some referrals also via primary care and social care.  Information about the service is often passed on through word of mouth and from workers within accommodation settings but the team will directly approach new arrivals by telephone to offer an appointment.    **Appointment times vary depending on needs** and whether the client is a family or a lone individual.  DNA rates addressed through active planning and reminders. |  | **Relationships with GP** practices vary but include sharing of space and resources, direct linking in with practice staff, and sometimes include working with a dedicated RESPOND GP.  **Expert migrant health MDT** brings together a range of health, social care, housing, education and other support services to support informed working and a holistic care response. Operates outside of standard care pathway with dedicated fast-track for specialist referrals.  Partnerships across the NHS, with local authority, Early Help, housing, education and welfare services, and the voluntary and community sector. |
| **Electronic and handheld health plan** follows an appointment and flags health needs and referrals to GPs, supports engagement with other services and stays with the family or individual as they are moved around the country. | |  | **Telephone interpretation** used, including when making appointment bookings.  **Family-based care approach** supported by informal childcare and play activities provided by HCA to support engagement with parents and allow for sensitive conversations.  **Trauma informed** care approach.  **Very flexible** approach to working across and between boroughs, sites, sectors, with tertiary services, and GP practices and with the complexity of needs. |  | **Regular** training and clinical psychology support through peer and specialist routes for staff to be able to reflect on their emotionally challenging work.  **Virtual RESPOND Advice and Guidance MDT** offered nationally to all services and locations for complex case planning, advice and guidance. |
| **Core activities delivered** by an Infection and Inclusion Health Practitioner (advanced nurse practitioner) and health care assistant taking a holistic and trauma-informed approach with additional capacity from consultants in paediatrics and infectious diseases providing specialist care. A broader team of clinicians offering immediate guidance to workers around complex cases and a dedicated administrative team supports appointment bookings. | |  |  |  |  |

RESPOND case example

### Case study 3.

Solentra is a non-profit organisation offering transcultural mental health care to people with a refugee or migrant background. They work across the three regions of Belgium, both individually with clients and in a systemic way with important figures, including extended family (sometimes in other countries), and professionals and support workers from across a person’s broader environment.

Their approach places human rights to good mental health and the United Nations Sustainable Development Goals^266^ as central priorities alongside their own model of care, known as PACCT (Psychiatry Assisting a Cultural diverse Community in creating healing Ties).^39^ PACCT takes a stepped care approach with heavy focus on daily stressors that may threaten psychological wellbeing and impact a persons’ coping resources. Step.1 of the approach focuses upon building local capacity to ensure that frontline responses to the individual are understanding of their situation and concerned with meeting their basic needs, such as housing, legal support, schooling, leisure activities and social contact; Step.2 uses community-based consultation to understand a person’s difficulties from within the cultural and migration frame of the individual (and their family) as well as the current societal conditions in which they are living. It aims to identify and articulate underlying assumptions, values and expectations held by each system or individual, to work with what clients themselves see as important, and to build a protective community; Step.3 involves one-to-one ethnotherapeutic consultations.

Their approach is flexible and can be outreach or clinic based. They have a core commitment to working with ethnotherapists or interpreters to deliver therapy in a client’s mother tongue, regardless of local language proficiency, and will aim to always work consistently with the same interpreter, interpreters that understand their framework and who often have shared experiences with clients, allowing them to perform as “mediators for therapy”.

Referrals are typically received through social workers, other psychologists, doctors and other health staff, school staff, language tutors, Guardians (workers or volunteers supporting integration of unaccompanied minors), support workers, and family. These referrers will be services (or family) with which Solentra will aim to work closely.

Recognising the challenges for broader professionals in communicating and cooperating across language and culture, they offer a range of online and in-person learning modules, intervision, and supervision. They deliver additional standalone psychosocial and mental health projects which have evolved in response to local need.

| **WHAT** | NON-PROFIT ORGANISATION TAKING A HUMAN RIGHTS APPROACH TO TRANSCULTURAL MENTAL HEALTH CARE | | | | |
| --- | --- | --- | --- | --- | --- |
| **WITH** | ASYLUM SEEKING AND REFUGEE CHILDREN AND YOUNG PEOPLE AND THEIR FAMILIES; SOME ADULTS; SERVICES IN CONTACT WITH ASYLUM SEEKERS AND REFUGEES | | | | |
| **WHERE** | OUTREACH INCLUDING RECEPTION CENTRES AND SCHOOLS, AND CLINIC-BASED, REGIONAL TEAMS ACROSS BELGIUM | | | | |
| **SOLENTRA**  **MODEL OF CARE** PACCT Model (Psychiatry Assisting the Cultural diverse Community in Creating healing Ties) | |  | **STEPPED CARE & PROJECTS** |  | **CAPACITY BUILDING**  Proactive and reactive – responding to need |
| **All projects draw on Solentra’s PACCT model** which frames transcultural mental health care around human rights and a broad ecological vision of health and takes a flexible, mobile and phased (stepped care) approach. PACCT focuses on working with a person’s protective environment, as a ‘whole system’. A primary focus is placed on daily stressors that impact a person’s coping resources and there is a heavy focus on learning from clients. | |  | **Helpdesk** phone line, with call-back function, as a first line discussion for care providers, including teachers and community workers working with refugees. |  | **Training and education courses** to support the knowledge, skills and attitude of those coming into contact with people with refugee backgrounds. Including: psychosocial support of refugee families, transcultural skills, detecting psychological problems and psychosocial education, and addressing resilience amongst migrants and refugees. |
|  |  |  | **Community-based consultation** (with schools, social workers or other support service, and usually the family) to form a protective community for a child. A form of training on the job the approach aims to enable all ‘players’ to understand a situation, be part of defining the issues, and part of defining the solutions. |  |  |
| **Multidisciplinary team** with a range of skills, experience and backgrounds (clinical, therapeutic and professional expertise, working in other countries, bi- and multi- lingual, shared culture or experiences with clients). | |  | **Ethnotherapeutic Consultation** referrals will be accepted where advice, training and community consultations have not been enough. Involve cross-cultural validated diagnostic instruments (where available), always with translators or intercultural mediators to facilitate therapy in the mother tongue of the client and/or ethnopsychologists. Flexibly delivered in recognition that people are in ‘survival mode’ and often facing many problems. Take a range of approaches including transcultural psychiatry, EMDR, psychotherapy, narrative exposure therapy, physicality and movement to express emotions and communicate. |  | **Training and peer review** provided to Public Centres for Social Welfare (and other services) to support services to understand a persons’ environment and deliver culturally sensitive work. This includes a focus on a person’s strengths and resilience; being aware and recognising the psychological issues refugees can face which impede the integration process and making appropriate referrals at the right time to the right person.  **Intervision and supervision** with own and staff of other services delivered flexibly in groups and individually. |
| **Translators and cultural intermediaries** always involved when considered necessary. | |  |  |  |  |
| **Independent** but collaborate with a regional mental health centre and deliver many projects in partnership, or for, state providers. | |  | **Other standalone projects** often evolve in response to need for example, social integration programmes, and a school-based youth mental health crisis group. |  | **E-Learning Academy** for culturally sensitive work: interactive modules including, building a relationship of trust, communication skills with unaccompanied minors, supporting well-being and the best interests of a child, community-based working, agency, and resilience. |

SOLENTRA case example 3

### Case study 4.

The Victorian Refugee Health Program, Australia is a nurse-led service providing holistic health assessments and care coordination to refugees and asylum seekers with complex needs. Service aims are set by the Victoria (state) Department of Health but delivery models, including the level of support and care that can be provided varies across 15 individual state services, influenced by a variety of factors including the structure of the local community health service, the size of a local refugee community, and the services urban/rural location.

Though not a gatekeeper of the GP or other health services, for more complex cases, large families, those with additional needs, and for asylum seekers outside of settlement support provision the Refugee Health Program will support people through all or most aspects of a Refugee Health Assessment, general health service navigation, and provide initial care management. This includes TB and other screening, identification of other physical or mental health or social needs, support with learning how to call an ambulance, how to attend the local hospital, and how to access GPs, optometrist, and dentists. They will provide crisis management, for new or previously discharged clients and if not already working closely with the GP, they will provide a report for GPs and any other referral services, highlighting a patient’s needs and investigations. Approaches are need-led, by the person or the family and may include wraparound support for those with more complex needs including social aspects, material needs and advocacy.

Teams are typically small but will always include nurses, sometimes, advanced nurse practitioners, and may include an immunisation nurse, practice nurse, social worker or care coordinator, and bi-cultural worker. Where teams are located within a large community health centre, there are typically strong internal (co-located) relationships with GPs, women’s health, counselling services, physiotherapists, speech therapists, dieticians, occupational therapists, early intervention and disability services, smoking cessation, family violence prevention, men’s behaviour change, drug and alcohol services, and with the practice reception and administrative staff. Though broader services are not refugee specific they sometimes share the same staff (who might work part time for different services) and will usually have substantial experience working with refugee clients.

They work closely with settlement case workers from where most referrals will come as well as youth services, schools, lawyers, and community and voluntary sector settlement organisations, charities and NGOs.

| **WHAT** | PHYSICAL AND MENTAL HEALTH ASSESSMENTS, CARE COORDINATION, HEALTH EDUCATION AND SPECIALIST REFERRALS | | | | |
| --- | --- | --- | --- | --- | --- |
| **WITH** | NEWLY ARRIVED ASYLUM SEEKING AND REFUGEE LONE ADULTS, FAMILIES AND CHILDREN, SOME ONGOING CARE FOR THOSE IN THE COMMUNITY REGARDLESS OF IMMIGRATION STATUS | | | | |
| **WHERE** | OUTREACH AND CLINIC BASED (USUALLY COMMUNITY HEALTH CENTRES) IN AREAS OF HIGH REFUGEE RESETTLEMENT, VICTORIA, AUSTRALIA | | | | |
| **VICTORIAN REFUGEE HEALTH PROGRAM** | |  | **CO-LOCATED SERVICES & PARTNERSHIPS** |  | **BROADER STRUCTURE** |
| **Typically, a nurse-led team** aiming to support increased access into health systems, health service navigation, initial assessment, some case management, health and care referrals, health advocacy and wider health capacity building. Work with newly arrived asylum seeking and refugee lone adults, families and children, some ongoing care for those in the community regardless of immigration status. Actively work to identify people who may have come in on a spousal visa often via established relationships with different communities.  Use of in-person and telephone interpreters. | |  | **Dispersed model** with 15 separate refugee health services and 60 funded Refugee Health Nurse positions across the state. Most are part of a Community Health model that is unique to Victoria. |  | **Nurse-led teams also support local capacity building** supporting culturally and population sensitive work both formally and informally, including training for workers in contact with refugees and asylum seeker, secondary consultation and support to other service providers, including GPs.  **National Refugee Health Service framework** is currently being developed. |
|  |  |  | **Direct referral** and close working with state settlement service who will refer individuals/families within a week to one month of arrival or sooner if health needs are flagged by IOM during pre-arrival screening. Health assessments will usually take place over a series of three or four visits. This extended period is seen as important for developing trust and relationships. |  |  |
| **Comprehensive health assessments** usually completed within one-month of a person arriving, encouraged over several appointments. Includes migration, social and medical history; women’s health; sexual health; psychosocial history including settlement stressors; physical examination; health screening; management and referral planning; and for children and adolescents, developmental, education and behavioural assessments. Some aspects include GP involvement. | |  | **Usually co-located** within a community health centre which are often seen as a community hub, typically located in lower socioeconomic areas, some with gym spaces, a pool, and community garden. One service is located within a tertiary hospital.  Co-located colleagues often also have high experience with refugee clients and can include counsellors, physiotherapists, occupational therapists, GPs, podiatrists, dentists and early intervention services. Some services receive satellite provision from specialist refugee health paediatricians and optometrists.  Services are usually well-linked with other organisations and a specialist trauma service (Foundation House). |  | **A state-wide facilitator** informs the Department of Health and policy about issues on the ground, supports workforce development with refugee health nurses and allied health professionals, offers particular support to the smaller more isolated/rural teams, and addresses capability within mainstream health services (e.g. to understand pre-arrival experiences and trauma-informed care). |
| **Team** varies by service but always includes a Refugee Health Nurse, often a bi-cultural worker, social worker or care coordinator. | |  |  |  | **Victorian Refugee Health Network** brings together the 15 services for collaboration, projects, development of resources, sharing knowledge, and national advocacy. Includes three **Refugee Health Fellows (**specialists in infectious diseases, child health and primary care) providing consultation to providers. |

VICTORIAN REFUGEE HEALTH PROGRAM case example 4

### Case study 5.

Bevan are a social enterprise providing “responsive” NHS General Practice and health and wellbeing services designed to meet the needs of people who are homeless, in unstable accommodation, or have come to Bradford and Leeds as refugees or to seek asylum. With a focus on holistic care, general practice is provided with the addition of onsite Arabic interpreters, includes a degree of advocacy, social referrals and letter writing, and is co-located with Bevan’s wellbeing centre which offers a range of informal and more structured wellbeing support.

A dedicated Young Person Worker works with young people, families, and schools, supporting access to mental health and recreational and social activities such as football and martial arts. A weekly young person’s well-being drop-in provides GP contact, health checks and wellbeing activities without appointment. A mental health lead supports mental health care planning and mental health and social referrals for adults.

An outreach ‘hotel’ team includes an immunisation nurse, nurse practitioner, health care assistants and a social prescriber, and provides clinical health checks and health screening, including some social history, offer of contraceptive counselling and contraception, catch-up vaccinations, some medical prescribing, and social prescribing to residents in temporary accommodation sites across the area. Their approach is to take a “holistic picture of people’s health” and can lead to a range of clinical and social referrals, including support with accessing community, social and educational activities. The team attempt to cover most residents in each site before moving onto another location. Assessments, including referrals are added to GP patient records.

Health and wellbeing activities take place in a dedicated wellbeing centre or outreach in communities, public spaces and temporary accommodation settings. Activities include a women’s group with a focus on women’s health and family planning delivered by a nurse, social prescriber, psychotherapist, GP and health advocates. Social prescribers and health advocates provide one-to-one support to explore people’s aspirations, support access to services and resources, and to support mental health. This includes a range of social, educational, and physical activity opportunities, and local service navigation.

There is heavy involvement of volunteer peer advocates supporting patients in the GP waiting room with a “welcoming face” and social support, help to complete NHS registration forms, make appointments, use the internet, and set up and operate the NHS App on people’s phones. Peer advocates also work alongside the different programmes, exploring people’s interests and signposting or supporting access to other groups and services. The broader staff team is multidisciplinary and also includes people with shared backgrounds to patients.

| **WHAT** | INCLUSION HEALTH GP AND HEALTH AND WELLBEING SERVICE | | | | |
| --- | --- | --- | --- | --- | --- |
| **WITH** | PEOPLE TYPICALLY EXCLUDED FROM CARE, INCLUDING REFUGEES AND ASYLUM-SEEKERS, SEX-WORKERS, AND PEOPLE WHO ARE HOMELESS | | | | |
| **WHERE** | A GP PRACTICE AND WELLBEING CENTRE, OUTREACH TO CONTINGENCY HOTELS, WEST YORKSHIRE, UK. | | | | |
| **BEVAN**  **HOLISTIC APPROACH**  GP service and health checks | |  | **HOLISTIC APPROACH**  Health and wellbeing service |  | **VOLUNTEERS (Usually former clients)**  Peer support |
| **Inclusion health GP surgery** providing NHS general practice care. Includes some advocacy, social referrals and letter writing. Some flexibility in appointments to accommodate walk-ins, alongside pre-booked and on-the-day appointments. In-house Arabic interpreters otherwise typically telephone interpretation for other languages. Active reception team liaise and refer to social prescribers, wellbeing services, and organise interpreter schedules in advance of appointments (where language needs known). Peer volunteers offer support to register with the practice. | |  | **Smart Health Inclusion Peer Advocates** providing peer support to enable socially excluded groups to improve digital skills and overcome barriers to access health and wellbeing services, including the internet, NHS App and online appointment booking. |  | **GP Reception welcome volunteers** offering a friendly introduction to the GP practice, supporting social connections and confidence in the waiting room, letting people know what services and activities are available, and delivering the **Smart Health Inclusion programme.** |
|  |  |  | **Young person’s project** dedicated young person’s worker providing one-to-one, family and community support, weekly well-being drop-in with GP contact, health checks and wellbeing activities without appointment. |  | **Group and one-to-one support** to support attendance and engagement in wellbeing, social prescribing and health information activities. |
| **‘Hotel’ temporary accommodation team** outreach support includes health screening, including mental health and TB and blood born viruses, catch-up immunisations, medical prescribing, and support to access and understand appropriate use of NHS services. Delivered by clinical and social prescribing team with some remote GP support available to workers. Social prescribers provide wellbeing activities and provide one-to-one support. Collaborate with community partners to deliver additional support and health information activities. | |  | **Social prescribing team** support social connections into the community and access to other services. Includes ESOL, college courses, arts activities, men’s woodwork, physical activities, support with making telephone calls, health and early years information sessions, volunteering. Constant evolution of new activities being tried. |  | **OTHER ASPECTS**  **Services developed** in consultation with service users  **Open kitchen** for all wellbeing visitors to make, tea, coffee, toast and lunch  **Some reimbursement** of travel costs  **ESOL classes and social workers co-located** during certain days of the week  **Food parcels** sometimes home-delivered |
|  |  |  | **Starting Well women’s group** offering group and one-to-one support including yoga, group-led and guided discussion in language specific groups on women’s health (contraception, smears, preconception care, mental health). Delivered by nurse, social prescriber, GP sexual health specialist, physiotherapist, peer volunteers. |  |  |
| **Mental health care planning** triage, on-the-day walk-ins, and case management providing mental health care planning and mental health and social referrals for adults. | |  | **Life in the UK** hotel based sessions focussed on supporting local and systems knowledge and understanding (e.g. when to call an ambulance, go to a pharmacy or go to the doctor and other UK-legal or social norms). Support with basic language, such as, “I need an interpreter”. |  |  |

BEVAN case example 5

#### Provision and staffing

##### Patients and clients

One service (case study 2) worked with asylum seekers, only, while all other services worked with people with a range of immigration statuses. One service (case study 5) also worked with people who were homeless or in insecure accommodation and sex workers; though there was some crossover between patient groups we focused specifically on their refugee and asylum seeker provision. All services worked with children, adults and families, one service (case study 3) worked heavily with unaccompanied minors and young lone males.

Four services worked predominantly with people who were newly arrived or within two-years of arriving within the local country. One service (case study 3) worked with people at any point in their settlement journey. Services stressed however that a needs-led approach typically governed the period of engagement and patients were often never formally discharged, with people told clearly how to get in touch and re-engage where needed.

##### Delivery setting, care provision and workers

Provision was delivered across a range of settings that included initial and contingency accommodation (4 services), health clinics (4 services), and community centres or spaces, including schools (3 services). Two of the services (case studies 4 and 5) based within health clinics were co-located with community, wellbeing and social support services and three services (case studies 2, 3, and 4) described occasional or initial home visits. A summary of offered provision is detailed in Table 4 and worker roles in Table 5.

| **PROVISION** | **DETAILS** |
| --- | --- |
| **Holistic Health Assessment & Health Screening** | Mental health; emotional and psychological wellbeing; experiences of torture and trauma; physical health; infectious diseases; sexual and reproductive health; child development; family functioning; safeguarding; migration/social/medical history; child behaviour; women’s health; oral and dental health.  Can lead to immunisations; prescribing; signposting; referrals.  Reports prepared for GPs and electronic and handheld health plan to stay with family/individual both (flagging health needs, mental health and social referrals). |
| **Health System Navigation** | Support with understanding and learning how to use local healthcare services; understanding rights to care; digital skills for using health service systems; locating and managing pharmacy/prescriptions; confidence building; managing expectations around health care, appointments and waiting times; physical support to access services (transport, accompanying); information and signposting to services; geographical mapping of services. |
| **Health Knowledge** | Health awareness/education sessions (various health conditions/needs including family planning, early years); ESOL for health. |
| **Health Advocacy** | Support with making/querying/chasing up appointments (telephone, online, in-person); drawing attention to urgent medical/dental care needs; challenging charging; bringing attention to concerns on individuals’ behalf; appointment reminders; support with understanding medical and Home Office correspondence; letter writing (multi-sector/social welfare/immigration). |
| **General Practice** | Inclusion health general practice; collaborating with or advising general practice; resource sharing; drop-in and walk-in provision; co-location with other services. |
| **Specialist consultation and Referrals** | Paediatrics, respiratory and infectious diseases, optometry, primary care, mental health (all in-house or satellite provision); rapid referrals; multi-sector referrals. |
| **Mental Health Care** | Transcultural approaches; ethnotherapeutic consultations; community-based consultation; mental health triage; crisis management and case work; dedicated young person care; trauma-informed care approaches; Eye Movement Desensitisation and Reprocessing (EMDR); psychotherapy; narrative exposure therapy. |
| **Social Prescribing, Social Support Assessments, Other health and wellbeing Provision** | Support identifying and connecting people with: community; social activities; hobbies; ESOL; education (school, college); arts activities; child play activities; social support services; employability support; faith and multicultural groups; volunteering opportunities; sport and physical activity opportunities; holiday activities. |
| **Dental Care** | In-house emergency care; dental referrals; help with making dental appointments. |
| **Other provision/practices** | Welcoming new arrivals; informal childcare; play activities; material goods. |
| **Capacity Building** | Training and awareness raising among other services; specialist supervision and intervision. |

##### Summary of care provision

###### Supporting workers

Services put substantial effort into supporting all staff and supporting staff resilience. This ranged from regular, formal and informal supervision (with clinical specialists or psychologists), reflective practice, and the use of designated meetings focussed on making sure staff were “okay” or to work through different cases and complex issues and share suggestions. One service had a WhatsApp group with consultants and GPs *on call* to frontline nurses, offering advice, sometimes mid-consultation. Elsewhere there were organised phone-ins from outreach staff to GPs, and an “open door policy” with staff encouraged to seek advice and support. There were also broader service network events bringing wider colleagues together as opportunities for professional development and to share experiences and knowledge.

| **CLINICAL** | **OTHER HEALTH WORKERS** | **OTHER WORKERS** |
| --- | --- | --- |
| Nurses; Advanced Nurse practitioners; Migrant Health Nurses; Inclusion Health Practitioners; Immunisation Nurses; Practice Nurses  Healthcare Assistants  GPs  Paramedics  Psychotherapists; Psychologists; Psychiatrists; Mental Health workers; Ethnotherapists  Occupational Therapists  Paediatricians  Infectious disease consultants | Social prescribers  Young Person’s Workers  Family Violence Workers  Health and Wellbeing Officers  Health advocates  Yoga Instructors | Peer Volunteers  Bi-cultural Workers; Intercultural Mediators  Interpreters  Social workers; Care Coordinators; Case workers  Front of house and reception teams; Administrative and Operational leads/teams.  Communications workers  Regional facilitators  Public health consultants |

##### Summary of workers
